# Restored somatosensation in individuals with lower limb loss improves gait, speed perception, and motor adaptation

**DOI:** 10.1101/2023.05.30.23290267

**Authors:** Daekyoo Kim, Ronald Triolo, Hamid Charkhkar

## Abstract

Lower limb loss is a significant insult to the body’s nervous and musculoskeletal systems. Despite technological advances in prosthesis design, artificial limbs are not yet integrated into the body’s physiological systems. Therefore, lower limb amputees (LLAs) experience lower balance confidence, higher fear of falls, and impaired gait mechanics compared to their able-bodied peers (ABs). Restoring sensations perceived as originating directly from the missing limb via implanted neural interfaces were shown to improve balance and performance in certain ambulatory tasks; however, the effects of such evoked sensations on neural circuitries involved in the locomotor activity are not well understood. In this work, we investigated the effects of plantar sensation elicited by peripheral nerve stimulation delivered by multi-contact nerve cuff electrodes on gait symmetry and stability, speed perception, and motor adaptation during walking. We found that restored plantar sensation increased stance time and propulsive force on the prosthetic side, improved gait symmetry, and yielded an enhanced perception of prosthetic limb movement. Most importantly, our results show the locomotor adaptation among LLAs with plantar sensation became similar to ABs. These findings suggest that our peripheral nerve-based approach to elicit plantar sensation directly affects central nervous pathways involved in locomotion and motor adaptation during walking. Our neuroprosthesis provided a unique model to investigate the role of somatosensation in the lower limb during walking and its effects on perceptual recalibration following a locomotor adaptation task. Furthermore, we demonstrated how plantar sensation in LLAs could effectively increase mobility, improve walking dynamics, and possibly reduce fall risks.

**One-Sentence Summary:** Neuroprosthesis stabilizes gait and improves speed perception and locomotor adaptation in individuals with lower limb loss.

## INTRODUCTION

Despite advances in prosthetic technology, current lower limb prostheses do not provide direct sensory feedback to the user. Consequently, individuals with lower limb loss need to rely solely on interpreting the pressures between their prosthetic sockets and residual limbs to learn indirectly about the interactions of their prostheses with the ground. This compromised sensory feedback partially plays a role in increased fall risks *(1)*, loss of confidence in the prosthesis *(2)*, and excessive reliance on the intact limb *(3)*. Asymmetrical walking, as often reported in lower limb amputees (LLAs) *(4)*, increases fatigue *(5)* and reduces postural stability during walking *(6)*. In addition, because the direct sensory link between the foot and the brain is missing and the perception of prosthesis-ground interactions is indirect, LLAs also have difficulties in adapting to changes in walking environments *(7–9*.

Plantar sensation and joint proprioception play a crucial role in continuously interacting with our surrounding environment while ambulatory *(10–12*, and provide information about changing surface characteristics to the central nervous system (CNS) aids in adapting to predictable perturbations such as walking on slippery surfaces or negotiating stairs. Input from plantar cutaneous afferents is integrated at different CNS nuclei during walking *(13)*, contributing to an accurate representation of the spatial and temporal movements of the limb *(14)*. Through the somatosensory feedback from lower limbs, we recalibrate limb speed perception and update the predictive internal model for controlling limb movement with our expected perceptual consequences.

For LLAs, the sensorimotor feedback loop is severely disrupted due to limb loss. Recent reports have demonstrated the feasibility of neural interface technology in restoring sensation perceived as originating from and co-located with the missing limb *(7, 15, 16)*. Sensations elicited by peripheral nerve stimulation in LLAs have exhibited modalities such as pressure, vibration, and perception of joint movement *(15–17)* referred to discrete locations on the missing limb. These reports also indicate improvements in balance *(18)*, ambulatory performance *(13)*, prosthesis weight perception *(19)*, and phantom pain *(20)* with appropriately modulated peripheral nerve stimulation. Despite such reports, it is not yet clear how elicited somatosensation from the missing foot affects motor function in the lower limb during ambulation. Furthermore, the effects of such elicited sensation on limb movement perception and the consequent impact on motor adaptation are largely unknown. Locomotion is inherently a sensorimotor task, and any elicited sensations from the lower limb are only effective if integrated into the motor circuitry.

The tactile sensation elicited by peripheral nerve stimulation suggests that areas associated with anatomical representations of the missing limb are activated in the primary sensorimotor cortex (S1) *(21)*. While multiple groups have investigated the neural pathways involved in the tactile sensation of hand, there has been less focus on determining the relationship between afferent inputs from the lower limb and higher neural circuitries, such as the spinal cord, cerebellum, and somatosensory cortex. Evidence from animal studies shows that the S1 processes afferent information from the foot and responds rapidly after the onset of perturbations suggesting a causal link between S1 area activation and motor adaptation and learning *(22–25)*. Hence, somatosensation elicited via peripheral neural stimulation may activate the same brain circuitries in LLAs and possibly drive motor adaptation and learning directly in a manner similar to individuals without limb loss.

In this work, we hypothesize that plantar sensation directly affects CNS pathways involved in limb movement perception, the ability to perceive changes in limb movements, and motor adaptation during walking. Our study presents a number of important observations. Firstly, we demonstrate that a sensory neuroprosthesis (SNP) improves the perception of limb movement. The SNP provides the user with elicited plantar sensations corresponding in location and intensity to applied loads to the prosthetic foot (Fig. 1). Secondly, we show that the changes in limb movement perception due to the SNP improves the biomechanics of walking. Finally, we show that the SNP enables LLAs to adapt to changing gait conditions imposed by the split-belt treadmill paradigm in a manner similar to able-bodied individuals (ABs). Our findings provide new evidence on how somatosensation contributes to locomotion and motor adaptation while walking. These results demonstrate how a neurally-integrated lower limb prosthesis can lead to increased mobility, improved walking dynamics, and possibly a reduction of fall risks among LLAs.

**Fig. 1.**
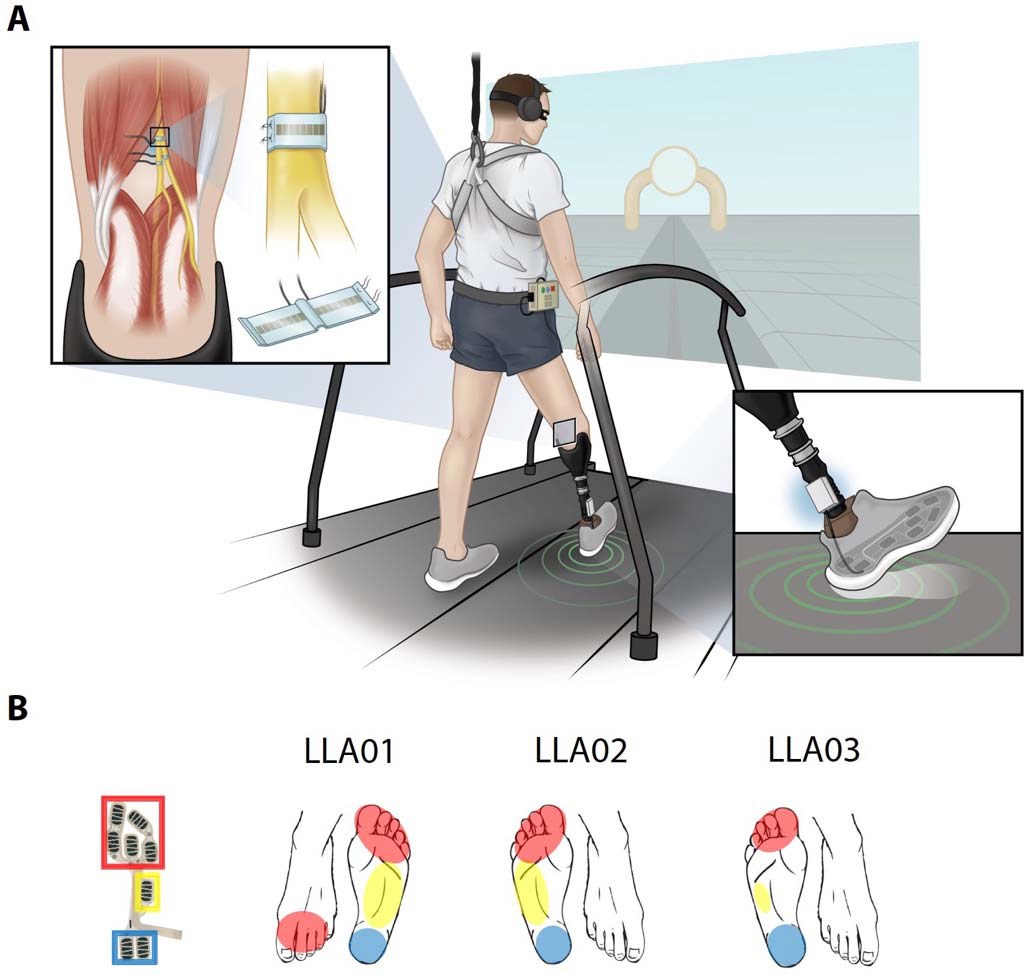
Neural interface technology and closed-loop somatosensory neuroprosthesis (SNP) **(A)** High-density C-FINEs were implanted around sciatic and/or tibial and peroneal nerves above the knee in three unilateral lower limb amputees (LLAs). The access to individual contacts within each cuff electrode was through percutaneous leads connected to an external stimulator. The SNP provided users with plantar pressure sensations as if they originated in the missing. Elicited sensations corresponded to foot-floor interactions in intensity and location while walking. **(B)** The perceived locations reported by pressing on the forefoot, midfoot, and rearfoot during walking are shown in red, yellow, and blue, respectively.

## RESULTS

### Restoring Somatosensation Improves Gait Symmetry and Stability

The artificially elicited sensation from the prosthetic limb was integrated into the intact neuromuscular control system, leading to immediate changes in gait. SNP users perceived sensations that matched in location and intensity of the plantar pressure under the prosthetic foot during walking, allowing them to easily perceive that the sensation was coming from the interaction of the prosthetic foot with the ground. To quantify the extent to which SNP improves walking, participants initially walked on a split-belt treadmill with belts moving at the same speed (Fig. 1A). As described in the Methods section, LLA participants performed this task with SNP in the active or inactive mode in a randomized order. We found the SNP active condition resulted in improvements in spatiotemporal gait symmetry during steady-state walking (Fig. 2). The prosthetic limb’s step length (SL) increased by an average of 7.55% for the SNP active versus the inactive condition (Fig. 2A, *P*<0.01). Average increases in SL for each participant were 7.85%, 7.40%, and 7.45% for LLA01, LLA02, and LLA03, respectively (Fig. S1A, B, & C). Similarly, the intact limb’s SL increased by an average of 15.87% for the SNP active condition (Fig. 2A, *P*<0.01). Average increases in intact limb SL for each participant were 16.20%, 14.38%, and 16.89% for LLA01, LLA02, and LLA03, respectively (Fig. S1A, B, & C). Notably, the SL symmetry changed from 105.90% to 98.34%, an average decrease of 7.13% (Fig. 2A, *P*<0.01): LLA01: 103.51% to 96.07% (7.18%), LLA02: 104.80% to 98.40% (6.11%), and LLA03: 109.38% to 100.56% (8.07%) (Fig. S1A, B, & C). For comparison, ABs showed an SL symmetry of 100.35% during the same task (Fig. 2A). Consistent with changes in SL, the prosthetic limb’s stance time (ST) increased by an average of 7.92% for the SNP active condition (Fig. 2B, *P*<0.01). The average increases in ST were 7.69%, 9.38%, and 6.89% for LLA01, LLA02, and LLA03, respectively (Fig. S2A, B, & C). The intact limb’s ST increased by an average of 8.28% for the SNP active condition (Fig. 2B, *P*<0.01): increases of 7.71%, 7.88%, and 9.25% for LLA01, LLA02, and LLA03, respectively (Fig. S2A, B, & C). Subsequently, the changes resulted in improvements in ST symmetry by an average of 8.95% (Fig. 2B, *P*<0.01) (LLA01: 9.27%, LLA02: 10.65%, & LLA03: 6.99%; Fig. S2A, B, & C). For comparison, ABs showed an ST symmetry of 100.06% (Fig. 2B).

**Fig. 2.**
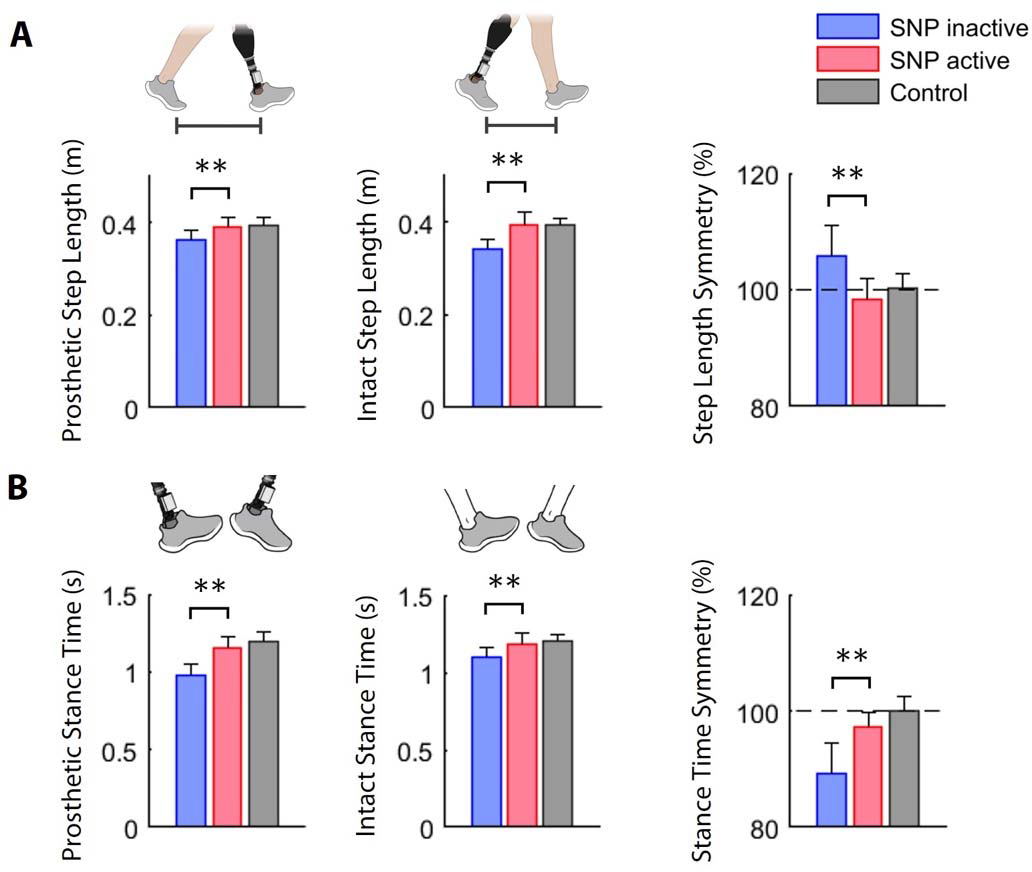
Spatiotemporal gait measures. **(A)** Step length (SL) from each limb and symmetry between limbs during the steady-state walking at 0.5 m/s. For the symmetry value, 100% indicates perfect symmetry, whereas values above 100 indicate longer prosthetic SL and hence more spatial asymmetry in the gait **(B)** Stance time (ST) from each limb and symmetry between limbs during walking. With a lower symmetry value, the prosthetic ST is shorter than the intact ST, and the gait is temporally more asymmetric. Bar graphs are mean values, and error bars are standard deviations. The asterisks indicate significant differences. \*\**P*<0.01.

We also found that the SNP increased ground reaction forces (GRFs) on the prosthetic limb. The peak propulsive force on the prosthetic limb increased by an average of 45.01% in the SNP active condition (Fig. 3A, *P*<0.01): increases of 27.74%, 51.79%, and 56.39% for LLA01, LLA02, and LLA03, respectively (Fig. S3B, E, & H), improving propulsive force symmetry by an average 48.96% (Fig. 3A, *P*<0.01) (LLA01: 19.25%, LLA02: 65.93%, & LLA03: 59.39%; Fig. S3B, E, & H). The peak braking force and symmetry remained unchanged for both SNP active and inactive conditions. In addition, the peak mediolateral force and symmetry remained unchanged for both SNP active and inactive conditions (Fig. S4A, B, & C) due to the simultaneous increases in mediolateral forces in both limbs. The peak vertical force remained constant in prosthetic and intact limbs regardless of the SNP condition (Fig. S4D, E, & F). However, the vertical force impulse on the prosthetic limb significantly increased by 6.99% due to increased stance duration on the prosthetic limb, thus improving the symmetry of the vertical force impulse between the prosthetic and intact limb by 6.22% (Fig. S4F).

**Fig. 3.**
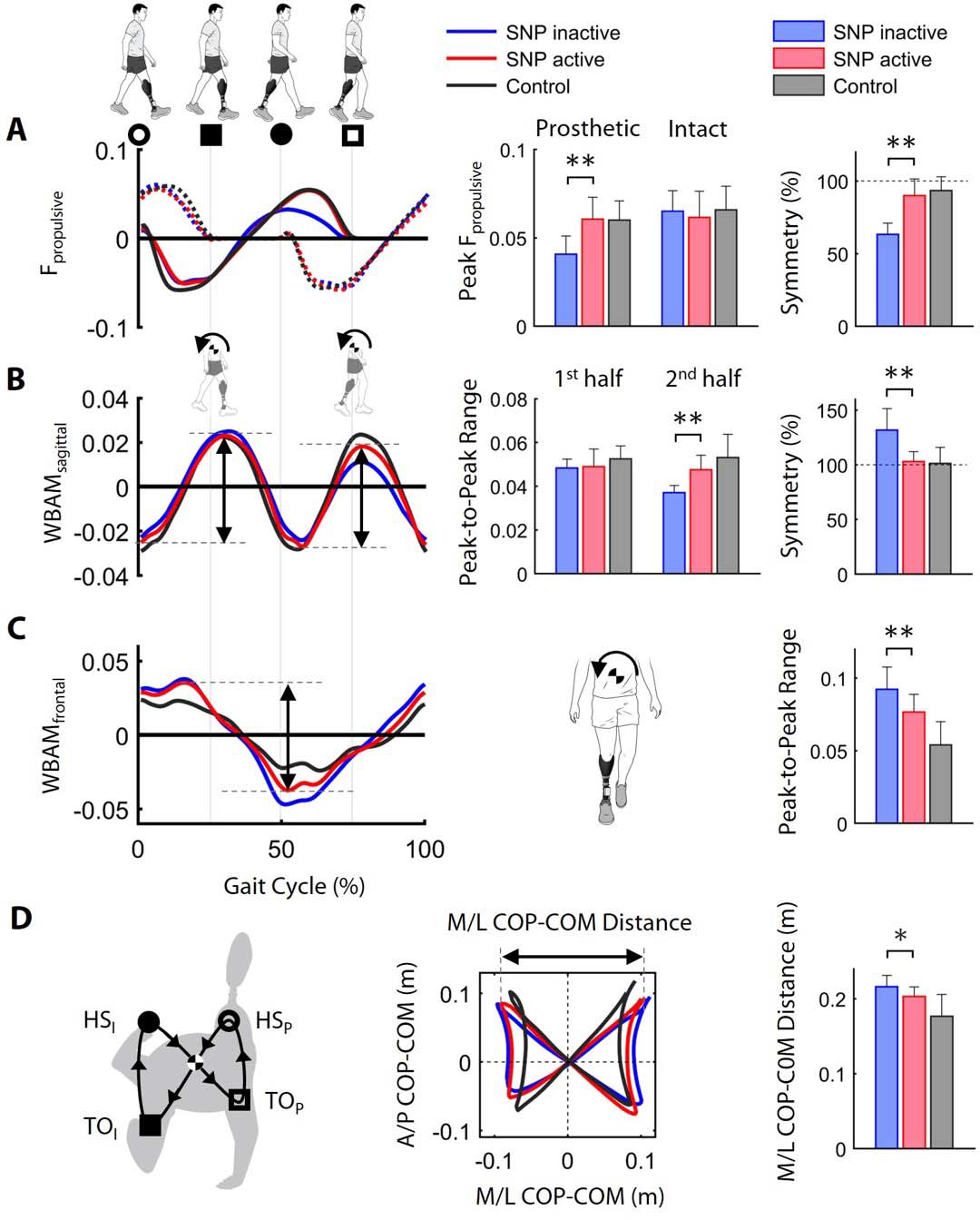
Dynamic gait stability measures. **(A)** Mean normalized ground reaction force (GRF) in the anterior (F_propulsive_) direction over the prosthetic limb gait cycle while walking at 0.5 m/s. Bar graphs show the peak F_propulsive_ from each limb and its symmetry between limbs. (B & C) Mean normalized whole-body angular momentum (WBAM) in **(B)** the sagittal plane (WBAM_sagittal_) and **(C)** the frontal plane (WBAM_frontal_) over the prosthetic limb gait cycle. In the sagittal plane, the positive WBAM represents a backward rotation of the body, whereas, in the frontal plane, the negative WBAM represents the body rotating toward the prosthetic side. The peak-to-peak range of WBAM is marked with dotted lines and arrows in the line graphs. The bar graphs in **(B)** show the mean ranges of WBAM_sagittal_ in the first and second half of the prosthetic limb gait cycle and its symmetry between both halves. The bar graph in **(C)** shows the mean range of WBAM_frontal_ over the gait cycle. **(D)** Butterfly pattern of COP profile relative to the body’s COM (i.e., COP-COM distance) measured during walking and the illustration of heel-strike (HS) and toe-off (TO) detection times (represented by circles and squares, respectively) for each limb. Solid and open symbols represent prosthetic and intact limb gait events, respectively. The direction of COP progression during the gait cycle is shown with black arrows. The line plot shows the distances between COP and COM as one series of x-y coordinates. The bar graph shows the mean COP-COM distance in the mediolateral (M/L) direction. The asterisks indicate significant differences. \*\**P*<0.01; \**P*<0.05. HS_P_: Prosthetic heel strike, TO_I_: Intact toe-off, HS_I_: Intact heel strike, & TO_P_: Prosthetic toe-off.

The whole-body angular momentum (WBAM) stabilized over the gait cycle in the SNP active condition. The range of sagittal-plane WBAM in the second half of the gait cycle (i.e., the intact stance phase) increased by an average of 25.34% in the SNP active condition (Fig. 3B, *P*<0.01) (LLA01: 53.52%, LLA02: 12.99%, & LLA03: 7.63%; Fig. S5A, C, & E). In addition, the range of frontal-plane WBAM was reduced by an average of 13.25% in the SNP active condition (Fig. 3C, *P*<0.01) (LLA01: 22.44%, LLA02: 10.40%, & LLA03: 1.59%; Fig. S3B, D, & F). The mediolateral center of pressure (M-L COP) was reduced by an average of 6.40% in the SNP active condition (Fig. 3D, *P*<0.01) (LE01: 10.23%, LE02: 10.35%, & LE04: 2.36%; Fig. S6A, B, & C). Despite changes in spatiotemporal gait measures, there were no changes in the ankle, knee, and hip joint angles for prosthetic and intact limbs during the stance phase of walking in LLAs, regardless of SNP condition (Fig. S7A-F).

### Enhanced Gait Speed Perception

To assess changes in limb speed perception, we designed a symmetry judgment task (SJT) (Fig. 4A). Consistent with standard psychophysical tests in sensory neuroscience, the SJT is a binary test to quantify the percentage of correct responses with and without SNP active. Participants performed the SJT to investigate whether SNP enhanced limb speed perception during walking, and to show how perceptual results were corroborated with actual biomechanical changes during walking. This highlights the suitability of SJT as a new task to investigate the relationship between the perceived and actual movement speed during ambulation. The belt speed on the intact side was kept constant, whereas it was changed on the prosthetic side in randomized order within the trial (See Methods section for more details). Verbal responses from the participants on whether the belts were at the same speed determined the “perceived speed,” while the center of mass velocity (COMV) was used to examine the actual speed symmetry between limbs (Fig. 4D). We found that LLAs with the SNP could more accurately match the speed of their prosthetic limb to that of the intact limb, indicating improved perceived speed (Fig. 4B). During the SJT, the perceived symmetry improved by, on average, 40.28% (Fig. 4C, *P*<0.01) in SNP active condition (LLA01: 50.01%, LLA02: 29.17%, and LLA03: 37.50%; Fig. S8A, D, & G). The LLAs with the SNP active also showed an improvement in the COMV symmetry by an average of 6.76% (Fig. 4E & F, *P*<0.01) (LLA01: 7.13%, LLA02: 7.34%, & LLA03: 5.15%; Fig. S9A, D, & G). The COMV symmetry measure supports the perceived symmetry results. Additionally, the time delay to verbal response was reduced by an average of 19.51% (Fig. 6A, *P*<0.01) when SNP was active (LLA01: 17.84%, LLA02: 24.05%, & LLA03: 28.42%; Fig. S10A, D, & G). The above results strongly suggest that our peripheral nerve-based approach to eliciting plantar sensation directly and positively affects central nervous pathways involved in gait symmetry and perception.

**Fig. 4.**
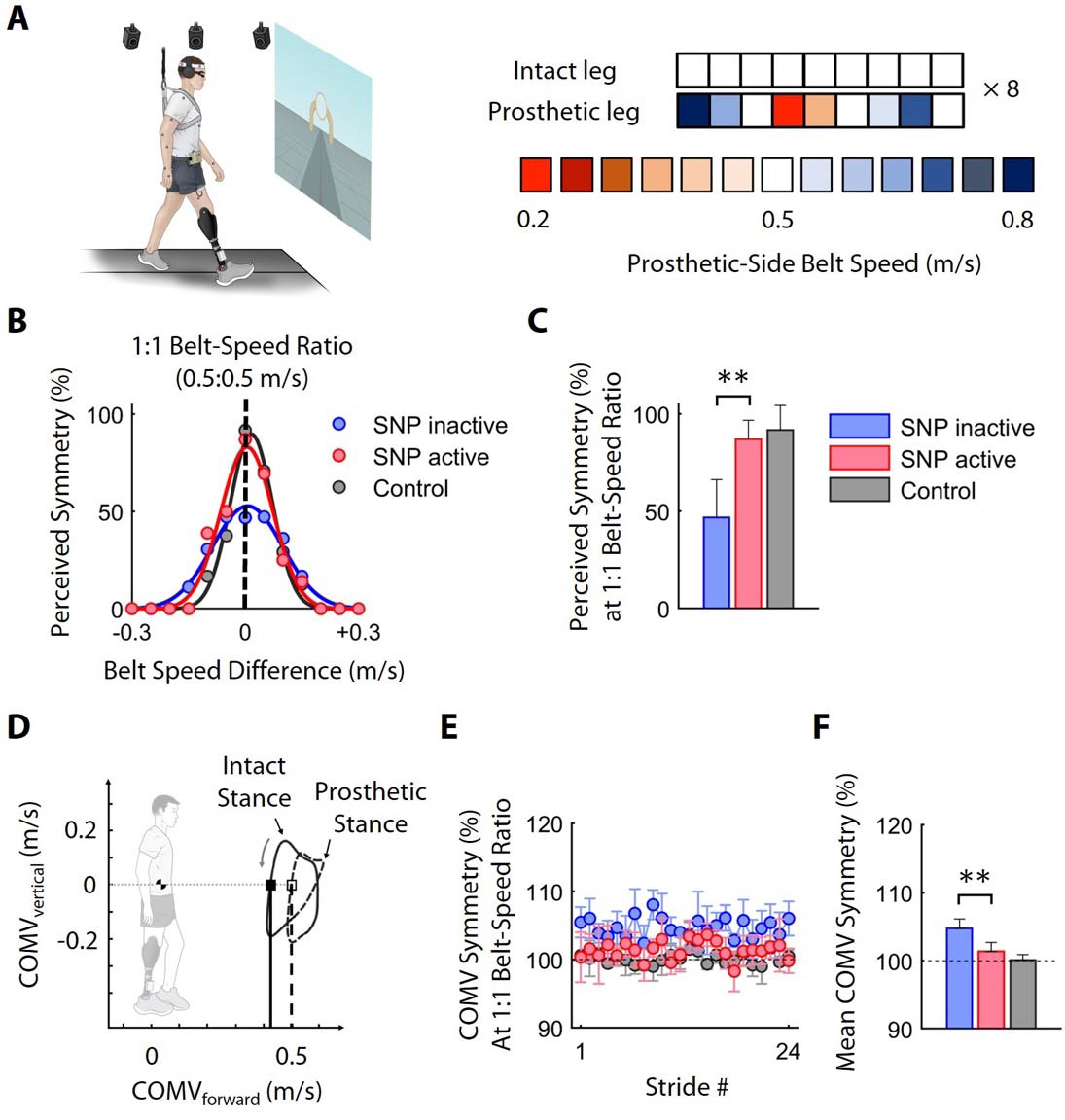
Perceived and actual gait symmetry measures. **(A)** The symmetry judgment task (SJT). Participants verbally expressed whether they perceived the speed of their prosthetic and intact limbs to be the same. The speed for the prosthetic-side belt was changed between 0.2 to 0.8 m/s at steps of 0.05 m/s (total of 13 levels) in a randomized order, while the intact-side belt was kept at 0.5 m/s. Participants wore dribble goggles to restrict their downward vision. Auditory cues from the treadmill motors were canceled via noise-cancellation headphones playing white noise. Participants wore safety harnesses on their chests, suspended from the ceiling at a non-weight-supporting level. **(B)** The mean perceived symmetry responses across all speeds. The x-axis shows speed differences from the base speed at 0.5 m/s. The black vertical dotted line indicates when belt speeds were the same. Gaussian curve fitting was used to define symmetry curves. **(C)** Percentage of perceived symmetry responses when belt speeds were the same. **(D)** The hodograph shows the COMV trace in vertical versus forward directions during a gait cycle. The trace moves in a counterclockwise path. For LLAs, when the vertical COMV is zero during the stance phase, the forward COMV on the intact side is lower than on the prosthetic side. **(E)** The COMV symmetry over 24 strides in which the belts had the same speed. **(F)** Mean COMV symmetry during SJT. In all plots error bars are the standard deviation. The asterisks indicate significant differences. \*\**P*<0.01.

### Effects of SNP on Locomotor Adaptation

In some situations, it has been shown that sensory feedback improves motor adaptation by repeating the same movement pattern, that is, the result of reinforcement learning. Here we confirmed whether LLAs with restored plantar sensation experienced perceptual changes similar to ABs in the context of split-belt adaptation. To determine the effect of the SNP on perceptual recalibration after split-belt walking, participants performed a motor adaptation task (MAT), where the prosthetic-side belt was driven to move at twice the speed of the intact-side belt. The SJT was performed pre- and post-MAT to characterize the effects of motor adaption on perceptional recalibration of walking speed (Fig. 5A). In the early MAT, the COMV symmetry was not statistically different between SNP active and inactive conditions (Fig. 5B & C), indicating that somatosensory feedback did not affect the initial adaptation in the task. Although LLAs appeared to adapt their limb speed to the fast belt as the MAT continued, the extent LLAs adapted over time depended on the SNP condition, with slower adaptation for SNP inactive condition. In the late MAT, the COMV symmetry improved for SNP active by 10.55% on average (Fig. 5B & C, *P*<0.01) (LLA01: 17.76%, LLA02: 6.06%, & LLA03: 8.23%, Fig. S9B, E, and H), indicating that sensory feedback increased the amount of adaptation to the fast belt. Interestingly, LLAs with SNP showed a similar adaptation to ABs in their walking during the MAT, suggesting the plantar sensation’s role in normalizing the motor adaptation in LLAs (Fig. 5B & C). The findings from the MAT provide preliminary evidence that plantar sensation has a direct role in locomotor learning, and our peripheral nerve-based technique in human amputees enabled us to probe this relationship in a systematic way.

**Fig. 5.**
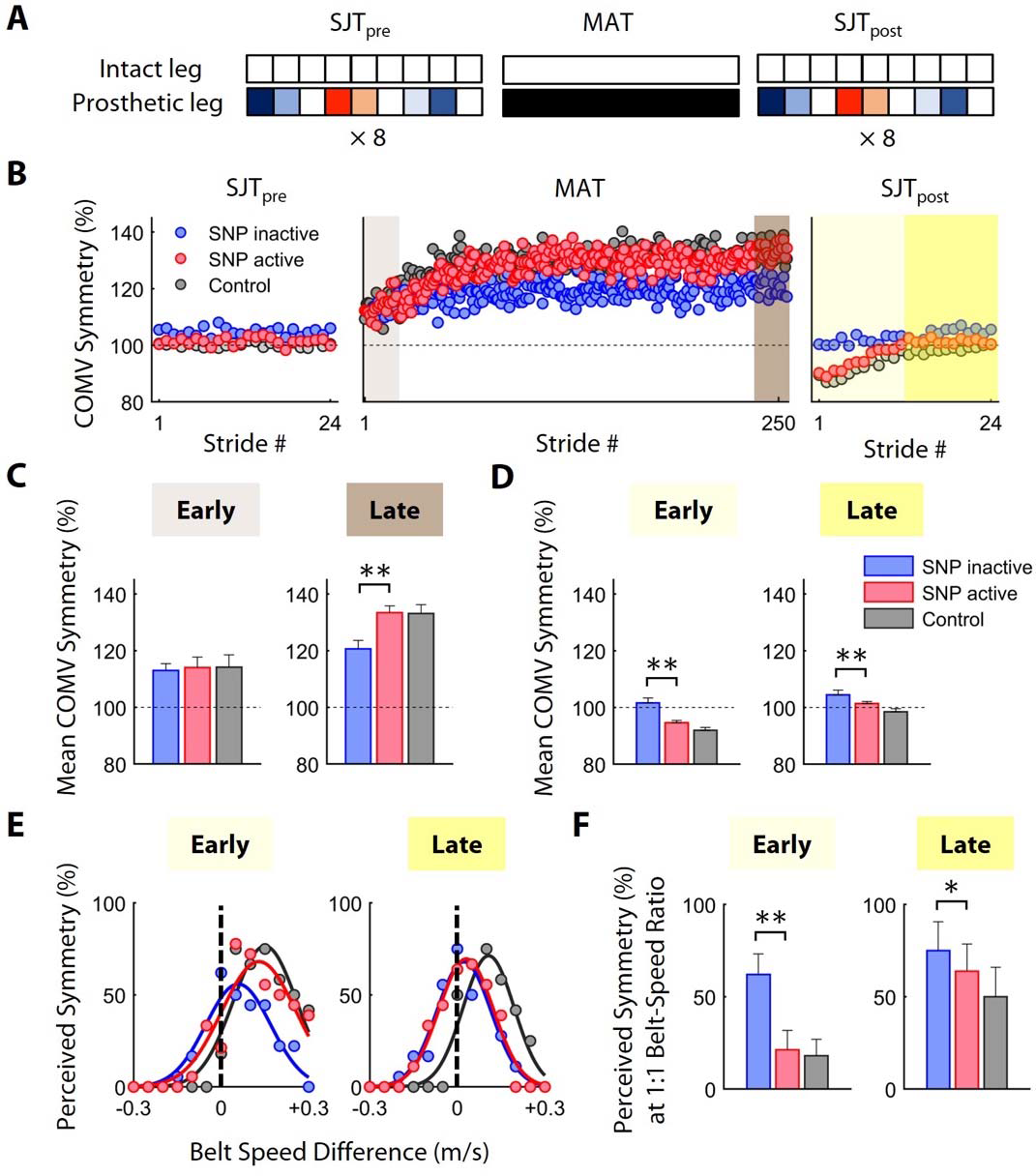
Motor and perceptual adaptation and aftereffects. **(A)** The motor adaptation task (MAT) was preceded and followed by SJT_pre_ and SJT_post_. During MAT, participants experienced a split-belt walking condition at a 2:1 belt-speed ratio (i.e., 1.0 m/s for the prosthetic side and 0.5 m/s for the intact side) for 10 minutes. The procedures for SJT_pre_ and SJT_post_ were identical to the SJT described in Figure 4. **(B)** Mean COMV symmetry over strides during SJT_pre_, MAT, and SJT_post_. For SJT_pre_ and SJT_post_, data are from strides when belt speeds were the same. **(C)** Mean adaptation in the COMV symmetry during the early MAT (i.e., first 20 strides, light brown) and the late MAT (i.e., last 20 strides, brown). **(D)** Mean aftereffects in the COMV symmetry during the early SJT_post_ (i.e., first 12 trials, light yellow) and the late SJT_post_ (i.e., the last 12 trials, yellow). **(E)** Mean aftereffects in the perceived symmetry across 13 belt-speed differences in the early SJT_post_ (i.e., first 36 trials, light yellow) and the late SJT_post_ (i.e., the last 36 trials, yellow). The black vertical dotted lines indicate when belt speeds were the same. **(F)** Mean aftereffects in the perceived symmetry at a 1:1 belt-speed ratio (i.e., zero belt-speed difference) in the early SJT_post_ (i.e., first 12 trials, light yellow) and the late SJT_post_ (i.e., the last 12 trials, yellow). In all bar graphs, error bars are the standard deviation. The asterisks indicate a significant difference. \*\**P*<0.01; \**P*<0.05.

To examine the effects of MAT on motor and perceptional recalibration, we performed another SJT following the MAT, i.e., SJT_post_. In the early SJT_post_, the COMV symmetry was decreased by an average of 6.98% in SNP active condition (Fig. 5D, *P*<0.01) (LLA01: 7.59%, LLA02: 6.18%, & LLA03: 6.98%, Fig. S9C, F, & I), while remaining unchanged in SNP inactive condition. In the late SJT_post_, although the difference in COMV symmetry in SNP active and inactive conditions became as small as 3.03%, they still remained significant (Fig. 5D, *P*<0.01). The COMV symmetry results confirm, support, and coincide with the subjective perceived symmetry measures. We also observed that LLAs with the SNP had similar perceptual recalibration to ABs in the early but not in the late SJT_post_ (Fig. 5E). Furthermore, LLAs with SNP demonstrated a robust adaptation (represented by a biased Gaussian curve in Fig. 5E) in the perception of their prosthetic versus intact limb speed. In the early SJT_post_, LLAs with SNP showed a significant adaptation aftereffect, resulting in a 65.74% decrease in perceived symmetry compared to the same group and condition pre-MAT (Fig. 5F, *P*<0.01): decreases of 71.87%, 56.95%, and 66.67% for LLA01, LLA02, and LLA03, respectively (Fig. S8B, E, & H), suggesting that the perception of limb speed was recalibrated such that the prosthetic limb (the limb that moved faster during MAT) felt slower after adaptation. On the other hand, the limb speed following MAT remained unaffected for SNP inactive condition (Fig. 5E & F), indicating no perceptual recalibration. Similar to the results observed from the SJT_pre_, the time delay to response in the early SJT_post_ was reduced by an average of 21.18% (Fig. 6B, *P*<0.01) in the SNP active condition (LLA01: 26.24%, LLA02: 13.38%, & LLA03: 23.32%, Fig. S10B, E, & H).

**Fig. 6.**
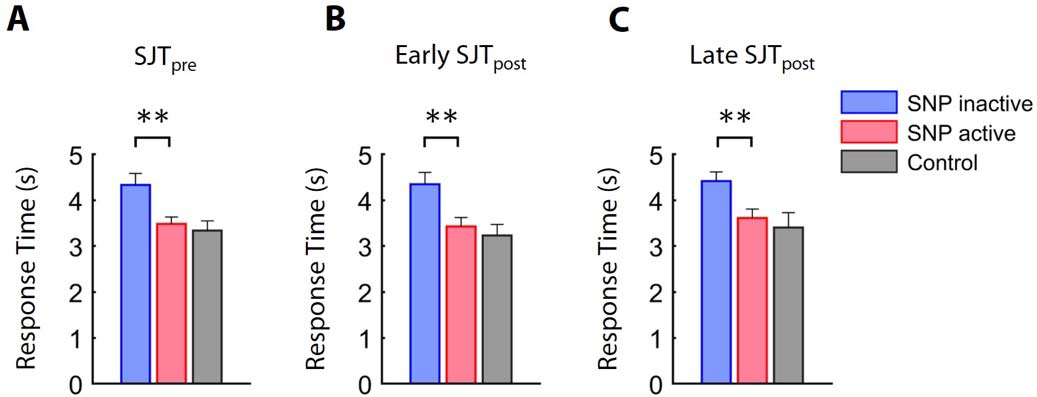
Response time during SJT. Given the same belt speed, the time the participants took to respond whether they perceived the belt speeds the same during **(A)** SJT_pre_, **(B)** early SJT_post_, and **(C)** late SJT_post_. Mean differences in response time are shown during SJT_pre_ (24 trials), early SJT_post_ (i.e., first 12 trials), and late SJT_post_ (i.e., the last 12 trials), respectively. Error bars are the standard deviation. The asterisks indicate a significant difference. \*\**P*<0.01.

## DISCUSSION

Somatosensation in the lower limb plays a crucial role in gait stability, symmetry, and control as it constantly informs motor planning and coordination. Our work shows that plantar sensation could have a meaningful functional impact among transtibial amputees, who constitute the majority of the LLA population *(26)*. It is reasonable to assume that similar or greater functional improvements could be achieved by SNP recipients with transfemoral amputation due to their higher gait stability needs. Furthermore, recent research indicates that the number of transtibial amputations among US veterans is increasing, while the number of transfemoral amputations is decreasing *(27)*. These findings suggest that despite the higher functional deficits experienced by transfemoral amputees, the growing demand for assistive technology for LLA is likely to come from the transtibial amputee population. Therefore, it is crucial to assess the effects of any new prosthetic technology on transtibial amputees to ensure the needs of prosthesis users within this population are met. Recent reports show how direct sensory feedback could improve temporal elements of gait (such as walking speed) for transfemoral amputees. Our study builds on such reports and provides a comprehensive analysis of the impact of elicited plantar sensation on spatiotemporal symmetry and whole-body dynamic balance, which are important and clinically relevant outcome measures of overall stability and movement dynamics that have not yet been studied. Above all, this study is unique and original in that it examines the effects of somatosensation on locomotor adaptation in LLAs which were not previously explored in *either* transtibial or transfemoral amputees. This is of particular interest because reduced or compromised somatosensation occurs as from of a variety of conditions, including highly prevalent peripheral neuropathy and dysvascular diseases such as diabetes that commonly result in transtibial amputation. Our results also provide insight into how plantar sensory feedback could improve motor learning, which could lead to new applications of neural interface technology for locomotor training-based rehabilitation that can maximize stability and safety in amputees and others with gait impairments resulting from sensorimotor impairment.

We demonstrated the effect of a lower limb SNP on the gait performance of unilateral transtibial amputees by developing a new walking speed perception task based on a split-belt walking adaptation paradigm *(28)* combined with psychophysical methods *(29–31)*. The experimental methodology was adapted from previously established motor adaptation paradigms utilized to assess ambulatory function in individuals with hemiplegia. This paradigm provides an experimental framework to understand how perceiving the interactions of the prosthesis with the environment during walking would affect motor learning and indicates how well the prosthesis is integrated into the body schema. It is directly related to real-world functional gait as we often change our speed during ambulation, and any mismatch between actual and perceived movement could increase the risk of falling. This paradigm is specifically relevant to maintaining gait stability during changing environmental conditions and exploring the robustness of the sensorimotor apparatus for safe and effective ambulation. Our results suggest that the plantar somatosensory feedback elicited via the SNP improved gait symmetry and stability, recalibrated the motor and perceptual response during walking, and updated the memory of the motor and perceptual recalibration after learning a new walking pattern. These findings indicate that the SNP directly affects locomotor neural circuitry in the CNS and reduces the discrepancy between the perceived and actual symmetry during walking.

We observed that the SNP reduced gait asymmetry by improving SL, ST, and GRFs. With the SNP active, the prosthetic foot pushed the ground stronger over a longer period, increasing the prosthetic foot’s forward propulsive force and ST, respectively. These observations suggest the elicited plantar sensations could partially compensate for the lack of prosthetic plantarflexion and increase the prosthetic ST to reduce the asymmetry between limbs. Longer ST on the prosthetic side led to an increase in the intact limb’s SL, thereby improving the symmetry between the prosthetic and intact SL. The increased propulsive force also indicates that plantar somatosensory feedback enhances the regulation of the prosthetic foot sole rolling over the treadmill belt, boosting the walking confidence of LLAs with the SNP. Elicited plantar sensations increased the forward propulsive force, but no change in vertical force was observed, as reported previously *(6, 13)*. This suggests that recipients of the SNP used the new sensory input to improve their gait mechanics to achieve a more regular gait pattern instead of developing a new, abnormal gait paradigm. Finally, the increased prosthetic forward propulsion increases work output from the prosthetic ankle during push-off *(32–34)*, potentially reducing the metabolic energy penalty for gait in LLAs *(35)*.

In addition to spatiotemporal gait improvements, the whole-body rotational dynamics changed with the application of the SNP. This is significant because it demonstrates the role of somatosensory feedback in maintaining dynamic stability to reduce fall risks and fall-related injuries in this population *(17)*. If not appropriately compensated, we found that the reduced prosthetic propulsion leads to alterations in whole-body movement patterns. This observation is consistent with reports following the use of powered ankle-foot prosthesis *(36)*, indicating somatosensation from the missing limb could provide higher propulsion and gait stability without the need for sophisticated, heavy, or expensive wearable robotic prostheses. Our results show that the SNP considerably increased prosthetic-side propulsion, intact limb SL, and, consequently, the range of sagittal-plane angular momentum in the second half of steps made with the prosthesis. These observations suggest that restored somatosensation on the prosthetic side compensates for the reduced propulsion in the step-to-step transition, contributes to symmetry in the inter-limb coordination during walking, and promotes forward and backward rotational stability. Although elicited sensations seem to affect sagittal-plane angular momentum, the mechanism for this change in whole-body rotation needs to be studied further. Recording hip and trunk muscle activity could help determine whether elicited sensations from the missing foot change any muscle activation patterns at proximal anatomical locations while walking and elucidate the mechanisms by which sagittal angular momentum is affected by the somatosensory inputs provided by the SNP.

Based on our results, the role of somatosensation is more dominant in maintaining mediolateral versus anterior-posterior gait stability. The level of awareness of prosthesis interactions with the environment was increased by the SNP, further highlighting the importance of adequate somatosensory contribution to adjusting the mediolateral COM position with the lateral border of the base of support (BOS). The SNP reduced the mediolateral distance between COP and COM (i.e., the distance between the vertical projection of the COM position and the lateral border of the BOS, see Fig. 3D) of LLAs over the gait cycle, bringing them to similar levels observed in ABs. This may be due to the SNP allowing LLAs to better control their BOS by adjusting the mediolateral COM state with appropriate strategies to reduce the mediolateral moment arm (i.e., position vector from COM to COP) in combination with higher forward propulsive force. The reduction in mediolateral COP-COM distance is the dominant mechanism for maintaining gait stability in the frontal plane *(37)*, contributing to the decrease in the range of whole-body angular momentum *(38, 39)*. Previous studies have shown that inadequate regulation of frontal plane angular momentum is indicative of poor balance control *(40)* and poor ability to adapt to perturbations *(41)*, particularly in populations with a high risk of falls *(42)*. Notably, transtibial amputees have difficulty regulating their angular momentum due to the functional loss of the ankle muscles *(43)*, which may explain their increased risk and fear of falling compared to ABs *(44)*. The restored plantar sensation significantly reduced the range of frontal-plane angular momentum, providing valuable insight into the biomechanical mechanisms that may detect and control the risks of falls in LLAs.

The improvement in limb speed perception with the SNP reveals an interconnect between proprioception and somatosensation. The symmetry judgment task (SJT) during ambulation allowed us to examine whether changes in the gait symmetry due to elicited planar sensations would affect the perception of walking symmetry between limbs. Without sensory feedback, participants relied on limited haptic information from residual limb-socket interactions, which were not sufficient to accurately perceive the limb movement during walking, as evidenced by poor perceived symmetry results during the SJT. However, with the SNP, our participants felt the change in the forward COMV in mid-stance with the prosthetic limb during walking. Hence, they were more aware of the speed differences between prosthetic and intact limbs. The improvement in the limb movement speed perception was possibly achieved with plantar sensation cues from the foot-floor interactions. Although the sensory perceptions elicited by the SNP were not directly related to proprioception, these results suggest that modulation of forward COMV due to the SNP provided the users with the feedback necessary to detect the timing of plantar sensations corresponding to the magnitude and location of prosthesis-floor interactions, from which they may have been able to infer proprioceptive-like information related to the ankle. In other terms, the elicited plantar sensations may have affected limb movement perception in an indirect way. During the SJT, the somatosensory feedback provided by the SNP enabled users to detect the changes in the belt speed and adjust their forward COMV accordingly. Thus, the SNP allowed LLAs to considerably reduce the discrepancy between the perceived and actual belt speeds and reliably discriminate subtle belt speed differences.

Our results from participants with transtibial limb loss show a lower forward COMV at mid-stance for the intact side compared to the prosthetic side, consistent with previous reports for both transtibial *(4)* and transfemoral *(45)* amputees. The improvement in the COMV with the SNP could be attributed to a lighter perception of prosthetic weight *(19)*, improved stability *(13, 18)*, and higher confidence in the prosthesis *(7)*. On the other hand, asymmetry of the forward COMV has been shown even when the prosthetic and intact limbs moved with identical kinematics *(46)*. Our results confirmed that plantar somatosensory feedback improves the symmetry of the forward COMV between prosthetic and intact limbs. With the SNP, the prosthetic ST increased, decreasing the forward COMV on the prosthetic side, which led to reducing COMV asymmetry.

We also investigated whether LLAs learned and retained a new walking pattern after a 10-min split-belt walking motor adaptation task (i.e., MAT). Without the SNP, the recalibration of limb speed perception was significantly reduced in the SJT_post_. This recalibration is an established perceptual aftereffect, such that the limb that moved faster during the MAT would feel as if it moved slower post-MAT *(47)*. In addition, the SNP helped users reach a perceptual recalibration similar to ABs. The perceptual recalibration effects are also supported by changes in the forward COMV symmetry measures, suggesting that the speed perception was affected by COMV changes during walking.

Our results demonstrate that plantar somatosensory feedback provides direct input to brain circuitries involved in perceptual recalibration during walking. Although sensorimotor learning is coordinated by the cerebellum *(48, 49)*, none of our LLAs had any reported cerebellar dysfunction. Prior work suggests that perceptual recalibration can occur even in patients with cerebellar deficit suggesting other brain areas, such as the posterior parietal cortex, might be involved in predicting sensory consequences of movement *(50)*. Without somatosensory feedback, our participants with lower limb loss were able to adapt their limb movements to the treadmill’s belt speed but could not update their perceived movement speeds. This suggests that in addition to pathways involved in motor adaptation, other areas in the brain, besides the cerebellum, might be involved in the perception calibration during walking in LLAs. The somatosensory feedback is received and processed in the somatosensory cortex *(51, 52)* and subsequently affects cortical motor control centers (such as the motor cortex) to revive the representation of the limb movement *(20, 53)*. Proprioceptive and somatosensory prediction errors then update the internal model, which consequently drives motor adaptation *(54, 55)*. Recently, Mathis et al. (2017) showed that the primary somatosensory cortex (S1) plays an essential role in motor adaptation in mice, which updates subsequent motor commands needed to reduce motor errors *(22)*. We found that LLAs without the SNP were similar to ABs in early adaptation (Fig. 5C Early) but different in motor (Fig. 5D Early) and perceptual (Fig. 5F Early) recalibration. This suggests that cutaneous input from the residual limb still played a role in motor adaptation; however, it was not enough to update the working memory of limb movement. Although lines of evidence, as seen in the human model, demonstrated the benefit of the SNP while walking on a treadmill *(56)*, over the ground *(17)*, and horizontal ladder *(13)*, this is the first report to examine the effects of elicited plantar sensation on motor adaptation. In this work, we were primarily concerned about whether motor adaptation was affected by amputation and could be normalized by the SNP. However, to understand the underlying mechanisms, in-depth motor-behavioral and imaging studies such as EEG recordings could help elucidate the relationship between SNP, activation of the sensory cortex, and its effects on motor adaptation.

Our work denotes several important functional implications of the SNP for LLAs, including achieving higher gait symmetry and stability, as well as the integration of the prosthesis with the neural circuitry responsible for locomotor adaptation. Although the emphasis during post-amputation rehabilitation is generally on achieving symmetric step length, partly because it is one of the most observable gait characteristics, attempts to achieve such symmetry may lead to other undesirable functional outcomes *(57, 58)*. The spatial measures such as step length asymmetry are highly variable, but subject-dependent, after lower limb loss. Therefore, a more robust measure to examine gait asymmetries is preferred. We argue that differences in COMV trajectories between prosthetic and intact legs may more accurately capture asymmetries in both magnitude and timing of loading between legs on a step-to-step basis. Most importantly, our results suggest that the COMV during the stance phase on the prosthetic limb is directly affected by the restored plantar sensation. Therefore, the symmetry of the forward COMV appears to reflect the actual kinematic behaviors exhibited by the lower limbs. Therefore, the sagittal COMV may be capable of detecting the timing of prosthetic loading during walking with a single, global variable that is also related to gait stability.

Although interfacing with the peripheral nerves following lower limb amputation has shown promising results in restoring somatosensation from the lower limb, our approach is uniquely different than the others. A detailed comparison of elicited sensations among different peripheral neural interfaces has been recently published *(59)*. Our perineural technology does not penetrate the nerve and, therefore, is particularly suitable for long-term use and applications outside of the laboratory. LLA01 received the surgically implanted SNP seven years prior to the time of this report, with consistent operation and stable sensory responses throughout. The data collection for this study lasted for approximately five months, during which time participants routinely visited our laboratory for other experiments following their implant surgery *(13, 15, 18, 60, 61)*. Throughout this and other previously reported studies, the implanted C-FINEs have demonstrated stable and reliable performance as evidenced by eliciting sensations in consistent locations related to the plantar surface of the foot of the missing limb. Furthermore, similar technology has been used in individuals with upper limb loss and spinal cord injury with no reported nerve damage or loss of response for over a decade *(62, 63)*, supporting the long-term stability and chronic viability of this neural interface. Moreover, our selective multi-contact nerve cuffs can significantly improve function with broader and less focused percept locations than reported in the previous studies *(7)*. We argue that smaller perceptual areas on the plantar surface of the foot might not be as crucial in improving gait mechanics as those on the palmar surface of the hand in enhancing grasp function. During gait, the sensory input from the plantar foot that contributes to the loading profile is primarily reliant on pressure sensing at a few broad regions, such as the toes, metatarsal areas, midfoot, and heel. Our technology can reliably and continuously elicit such sensations. Additionally, there is a similarity in reported modalities and intensities of sensations between our approach and the intraneural method. Both approaches reported qualities such as touch, pressure, and tingling that can be modulated over a useful dynamic range in response to input from physical sensors *(15, 17, 59)*. Most importantly, our findings on improvements in gait biomechanics have not been reported before. In this study, we report on improvements in gait symmetry and stability, and motor adaptation during walking. In contrast, the previously published work reported increased gait speed and improved gait efficiency (i.e., reduced metabolic cost).

We acknowledge that our study had certain limitations. Only three LLAs who received our implantable SNP participated in the current study. Therefore, data from a larger population, including participants with transtibial and transfemoral amputations, must be collected before generalizing the reported positive effects of restoring sensory feedback to LLAs. Because we used a new task paradigm, results need to be verified in a larger group of people with a more diverse demographic of age and sex with difficulty in walking due to various amputation etiologies.

## MATERIALS AND METHODS

### Research participants

Six able-bodied volunteers (AB01-AB06), three women and three men, served as a control group. On average, participants were 23 ± 3 years old (mean ± standard deviation) and 1.76 ± 0.07 m tall (See Supplemental Table S1 for more detailed demographics). Three volunteers with unilateral below-knee (transtibial) limb loss were also enrolled, received implanted multi-contact nerve cuff electrodes and associated components, and performed the study procedures (LLA01-03). All LLAs were male, with a mean age of 62 ± 7 years and a height of 1.75 ± 0.08 meters. None of the LLAs had a medical history of neuropathy, nor did they experience significant pain in their residual limbs. All LLAs were regular prosthesis users and wore their clinically-prescribed personal prostheses for all experiments. Two LLAs wore energy-storage-and-return prostheses, and one wore an actively powered ankle prosthesis (Ottobock Empower), which was unpowered and thereby locked at a neutral angle during experiments. The Louis Stokes Cleveland Veterans Affairs Medical Center Institutional Review Board approved all experimental procedures. Portions of this study involving the SNP were conducted under an Investigational Device Exemption obtained from the United States Food and Drug Administration (IDE G110043). All participants gave their written informed consent prior to any research-related activities, which were designed in accordance with the relevant human subject protection guidelines and regulations.

### Sensory Neuroprosthesis (SNP)

The SNP consisted of implanted components, an instrumented prosthesis, and an external controller. The implanted components included 16-contact composite flat interface nerve cuff electrodes (C-FINEs), which were connected to percutaneous leads exiting the body at the anterior mid-thigh (Fig. 1A). Each LLA received three C-FINEs surgically implanted around the sciatic and/or its main branches (tibial and common peroneal nerves). The external controller was connected to the implanted C-FINEs via the percutaneous leads. The delivery of electrical currents to the implanted C-FINEs elicited tactile plantar sensations perceived as if they originated from and were co-located on the missing foot. The external controller was placed in a fanny pack carried around each participant’s waist while walking (Fig. 1A).

The stimulation was delivered in a series of charge-balanced cathodic-first current pulses with the pulse amplitude (PA) ranging between 0.8 – 2 mA, and pulse width (PW) of 0 – 255 ms. The inter-pulse interval was always kept at 50 Hz, consistent with prior reports *(13, 18)*. The selected contacts for each C-FINE and their associated PAs were chosen based on threshold/mapping experiments described in Charkhkar et al. (2018) *(15)*, which consisted of compiling the verbal reports from the participants describing the location and modality of the elicited sensations, numerically rating perceived intensities, and repeatedly capturing the size and referred positions of the percepts referred to the missing or residual limbs by drawing on a digital tablet. The C-FINE contacts for SNP were then selected based on percepts reported in plantar regions of interest (e.g., heel, midfoot, and toes or metatarsal areas). For the implementation of SNP, the intensity of each elicited percept was linearly modulated while walking by changing the PW value based on readings from the insole pressure sensors. The full description of the mapping between the sensor and the stimulation output was given in Charkhkar et al. (2020) *(18)* and consisted of completely unloading the foot for baseline and full body weight on the region of interest (heel, midfoot, and toe) for maximal values to calibrate the interactive system.

Each user’s personal prosthesis was instrumented by adding a thin film force-sensing insole (IEE S.A., Bissen, Luxemburg) which was powered by a custom-made circuit board housed in a lightweight 3D printed enclosure attached to the pylon. The circuit board (i.e., the wireless sensor module) also transmitted the force data from each of the eight sensors embedded in the insole to the external controller. The external controller was programmed to deliver electrical stimulation within a safe range based on received force data from the instrumented prosthesis. The perceived intensity and location of the elicited sensation were adjusted by modulating the stimulation pulse width and the C-FINE contact, respectively. The details of surgical procedures, implantation techniques, neural interface technology, sensory characterization, and SNP setup have been reported in our previous work *(15, 18)*.

### Experimental Setup and Data Collection

A split-belt instrumented treadmill (R-Mill, ForceLink, The Netherlands) placed in a virtual reality (VR) facility (V-Gait, Motek Medical, the Netherlands) was used for all walking experiments. Each belt was driven by an independent motor. For simplicity, when two belts were driven at the same or different speeds, we refer to them as “tied belts” and “split belts,” respectively. The speed control was achieved via the system D-Flow software (version 3.16.2, Motek Medical, Amsterdam, the Netherlands). Force data were recorded from the two force plates embedded in the split-belt treadmill at a 1,000 Hz sampling frequency. Kinematic data was recorded via a full-body model (Plug-In Gait Marker Set). The marker trajectories were tracked using a 16-camera motion capture system (VICON, Oxford, UK) operating at 100 Hz.

### Experimental protocols

The first part of the experiment session served as a baseline. During the baseline, participants walked on tied belts for two minutes, where speed was kept constant at 0.5 m/s. Following the baseline, the experiment session consisted of the following phases: SJT_pre_, MAT, and SJT_post_ (Fig. 5A). The SJT was developed based on previously established psychophysical approaches to measure perception simultaneity *(64)*. During SJT, the treadmill belt under the prosthetic side was varied within 0.2 – 0.8 m/s at steps of 0.05 m/s, whereas the other belt was kept at the constant speed of 0.5 m/s the entire time (Fig. 4A). The 13 different speeds, ranging from 60% slower to 60% faster than the intact limb belt speed, occurred in a randomized order. Participants were asked to verbally announce whether they perceived both limbs at the same speed. The speed change happened every 6-10 seconds, depending on the participant’s response time. The total number of speed changes within the SJT was 72, which included four repetitions for each of the 12 split-belt speeds and 24 repetitions of the tied belts. During MAT, participants experienced a 2:1 (1.0 m/s:0.5 m/s) belt speed perturbation for 10 minutes (Fig. 5A). The faster belt speed always happened on the prosthetic side. The SJT_post_ condition was identical to SJT_pre_ and performed to determine any motor and perceptual changes following MAT. Although the speeds within the SJT blocks were randomized within each experiment, the order of speed blocks was kept the same between SJT_pre_ and SJT_post_ for each session to determine the aftereffects of MAT in both motor and perceptual domains. Due to the availability of participants, LLA01, LLA02, and LLA03 performed eight, six, and four experimental sessions, respectively. The experiments with LLAs lasted approximately five months, during which time SNP consistently elicited stable percepts with no change in reported locations, modalities, and intensities over time. The starting order of the SNP condition was randomized between participants, and they each performed equal sessions with the SNP active and inactive. ABs performed a single session only.

In all experiments, participants were positioned in the middle of the treadmill with one leg on each belt and wore a safety harness suspended from the ceiling (Fig. 4A). The safety harness did not support body weight and did not restrict their walking. Participants were blind to the timing and order of the belt speed changes. They were instructed not to hold onto the handrails on the side of the treadmill while walking unless they felt unbalanced. They wore dribble goggles to refrain from looking down at the belts or their feet and were instructed to look straight at the VR screen in front of the treadmill. Any auditory cues due to treadmill motors were canceled via noise-cancellation headphones (Sony WF-1000X, New York, NY, USA) playing white noise.

### Data analysis and outcome measures

Marker trajectories were labeled, and gaps were filled using Vicon Nexus 1.8.5 (Oxford Metrics). We filtered marker and force data with a zero-phase lag fourth-order Butterworth low-pass filter with a 5-9 Hz cut-off frequency selected by a residual analysis *(65)*. A subject-specific, 15-segment rigid body model, including head, forearms, upper arms, hands, trunk, pelvis, thighs, shanks, and feet, was created in Visual3D (C-Motion, Inc, Germantown, MD). The body segment parameters (mass, COM, and radius of gyration ratios) reported by De Leva (1996) were used to find the location and velocity of the whole-body COM *(66)*. The single force-plate center-of-pressure (COP) was obtained by scaling the COP position of each force plate with the magnitude of its respective GRF. The COP represents a weighted average of the pressures of the feet area in contact with the ground. Processed kinematic and kinetic data were exported as ASCII files and then imported into MATLAB to compute outcome measures.

We first measured two spatiotemporal gait parameters: step length (SL, the absolute difference in anteroposterior distance between the heel marker of each limb at the heel strike over a gait cycle) and stance time (ST, the time that the foot is on the ground between a heel-strike to the following toe-off of the same foot). We then calculated SL and ST symmetries as the percentage of the prosthetic to the intact SL and ST, respectively. To examine the effect of plantar sensation on limb loading response, we analyzed the GRF peaks and impulses at each limb, normalized by body weight (N). The GRF symmetry was defined as the percentage of the ratio of the prosthetic to the intact limb’s forces. The dynamic gait stability of LLAs was examined by analyzing the whole-body angular momentum (WBAM) about the body’s COM normalized by body mass (kg), height (m), and walking speed (m/s) *(42)*. The peak-to-peak range of WBAM in the sagittal and frontal planes of motion was subsequently calculated and reported *(6, 67)*. Furthermore, we analyzed the mediolateral distance between the COP and COM (i.e., external moment arm of the GRF in the frontal plane) to quantify the time rate of change in the frontal-plane WBAM. *(68)*. We measured the peak angles at the ankle, knee, and hip joints in the sagittal plane over the gait cycle to determine any changes in joint kinematics due to the SNP

We calculated the perception symmetry as a function of belt speed differences by fitting each participant’s ‘symmetric’ responses to a psychometric curve (i.e., Gaussian curve limited to 100%) during SJTs. Only psychometric curves with goodness of fit (R^2^) values above 0.5 were accepted for statistical analysis *(64)*. A single measure was extracted from the psychometric curve: the perceived symmetry at a 1:1 belt speed ratio (i.e., zero difference between belt speeds). This represents the percentage of true positives when participants correctly responded “Symmetry” when the belt speeds were actually the same. In all symmetry measures across experiments, a value of 100 indicated perfect symmetry. The symmetry value over 100 indicated a greater perceived speed on the prosthetic side than on the intact side and vice versa.

To corroborate the limb movement perception results with a comprehensive biomechanical measure during SJTs and MAT, we calculated the forward COMV at the moment the vertical COMV was zero during the stance period on each limb (Fig. 4D). The symmetry of the COMV was computed as the percentage of the ratio of the forward COMV during the prosthetic stance phase to the one during the intact stance phase *(46)*. COMV symmetry was previously utilized to account for the normalized speed difference performed by each limb on the body’s COM in individuals with lower limb loss and exhibited robust performance and consistency in the inverted pendulum model for amputee gait *(69)*.

### Statistical analysis

All data were exported and processed in MATLAB (R2021b, MathWorks, Natick, MA, USA) and reported as mean values ± SD. The normality of data distributions was verified using the Kolmogorov-Smirnov normality test. For the baseline, SJT_pre_, and MAT, a paired samples t-test was used to compare the outcome measures between SNP conditions (i.e., active or inactive). To determine the effects of SNP on motor adaptation, a two-way repeated measures ANOVA was conducted to compare the outcome measures for each SNP condition and task (i.e., SJT_pre_, MAT, and SJT_post_). Pair-wise post-hoc comparisons were performed using a Bonferroni adjustment. Significance was set at α < 0.05.

## Supplementary Materials

Table S1

Figs. S1 to S10

## Data Availability

All data needed to evaluate the conclusions in the paper are present in the paper or the Supplementary Materials.

## Acknowledgments

This manuscript has completed the review process in Science Robotics. We thank all our participants, especially SNP recipients for their dedication and time. We also thank Melissa Schmitt, RN, Alexandra Hutchison, LPN, Aarika Sheehan, DPT, Clay Kelly, MD, and Gilles Pinault, MD for their assistance with clinical and regulatory aspects of this study.

## Author contributions

DK and HC conceived the study design and planned the experiments. DK and HC conducted the experiments and initial discussions of the results and manuscript content. DK performed the data analysis. DK drafted the manuscript with contributions from HC and RT. HC and RT supervised the project. All authors contributed to design development, data interpretation, and manuscript editing.

