## Supplementary Tables and Figures for "Restored somatosensation in individuals with lower limb loss improves gait, speed perception, and motor adaptation"

Daekyoo Kim et al.

**This PDF file includes:**

Table S1

Figs. S1 to S10

**Table S1. Demographics and anthropometric information for participant characteristics enrolled in the study.**

| **SNP users** | **Age (yrs)** | **Height (m)** | **Gender** | **Affected Side** |
| --- | --- | --- | --- | --- |
| **LLA01** | 66-70 | 1.73 | M | L |
| **LLA02** | 56-60 | 1.68 | M | R |
| **LLA03** | 56-60 | 1.85 | M | R |
| **AB controls** | **Age (yrs)** | **Height (m)** | **Gender** | **Dominant Side** |
| **AB01** | 21-25 | 1.66 | F | R |
| **AB02** | 21-25 | 1.85 | M | R |
| **AB03** | 21-25 | 1.84 | M | R |
| **AB04** | 26-30 | 1.78 | M | R |
| **AB05** | 21-25 | 1.77 | F | R |
| **AB06** | 16-20 | 1.69 | F | R |

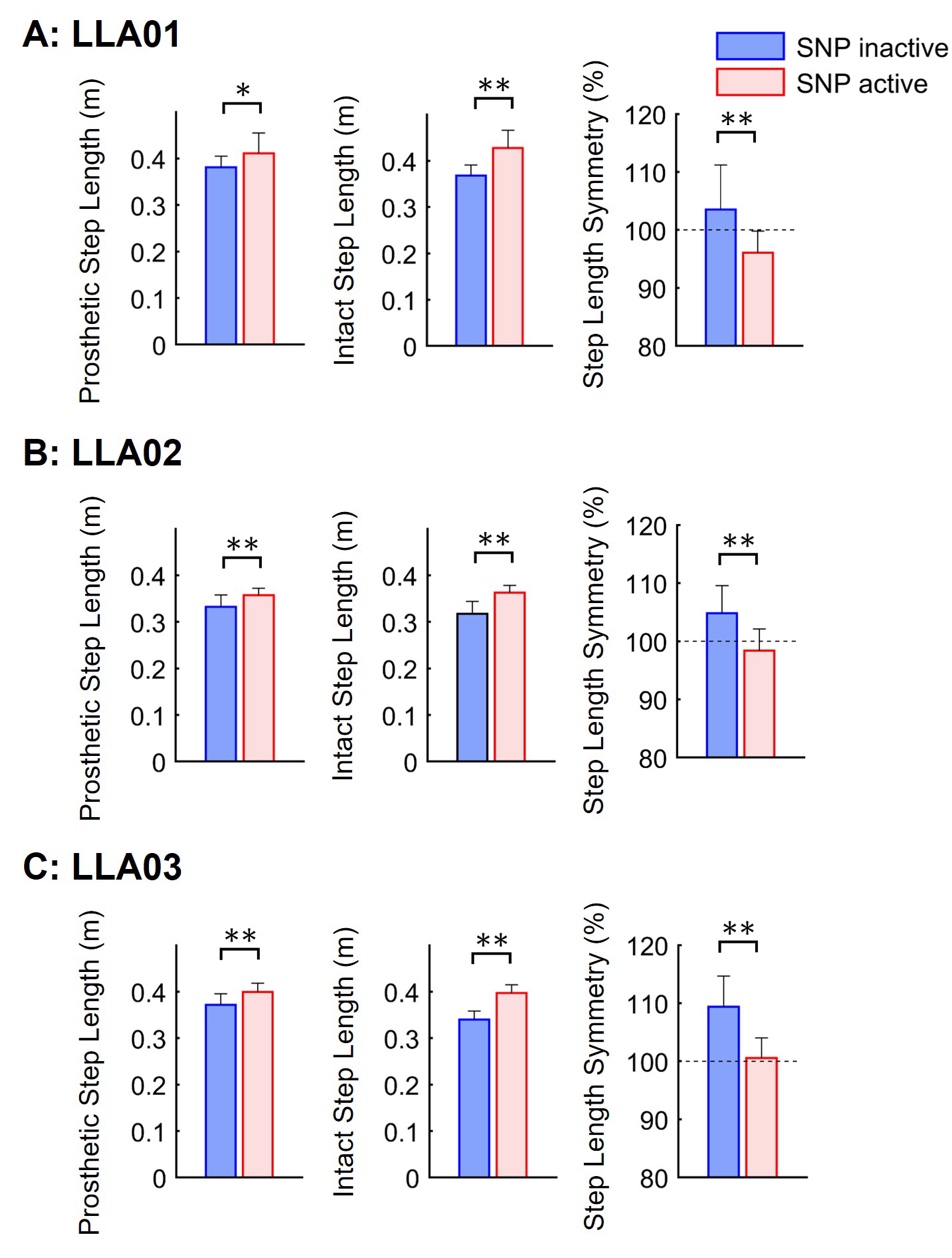

**Fig. S1. Individual SNP users’ step length (SL) measures. Related to Fig. 2A.** Mean SL from each limb and symmetry between limbs during walking for LLA01 **(A)**, LLA02 **(B)**, and LLA03 **(C)**, respectively. Error bars indicate the standard deviation. The asterisk indicates significant differences. ***P*<0.01; **P*<0.05.

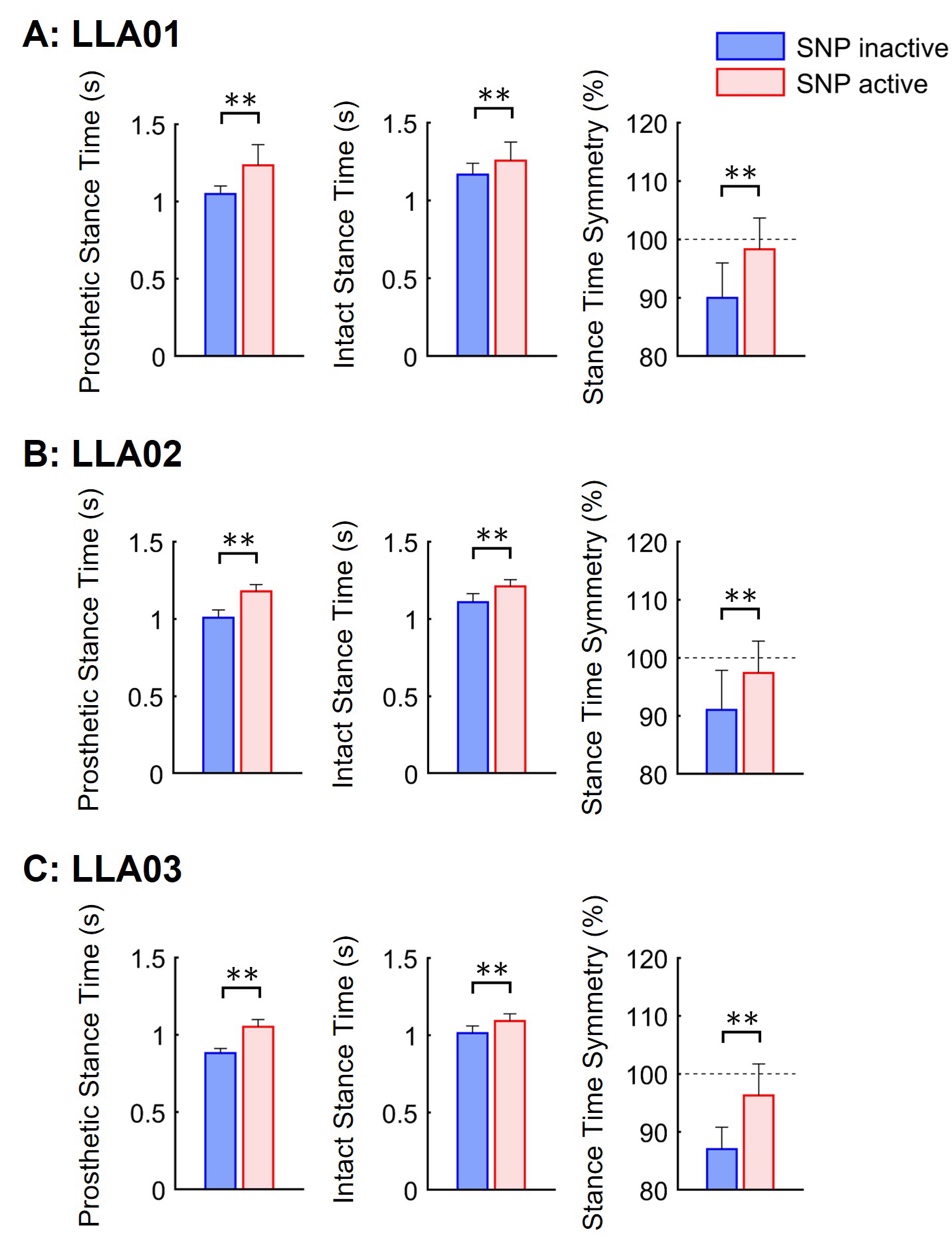

**Fig. S2. Individual SNP users’ stance time (ST) measures. Related to Fig. 2B.** Mean ST from each limb and symmetry between limbs for LLA01 **(A)**, LLA02 **(B)**, and LLA03 **(C)**, respectively. Error bars indicate the standard deviation. The asterisk indicates significant differences. ***P*<0.01.

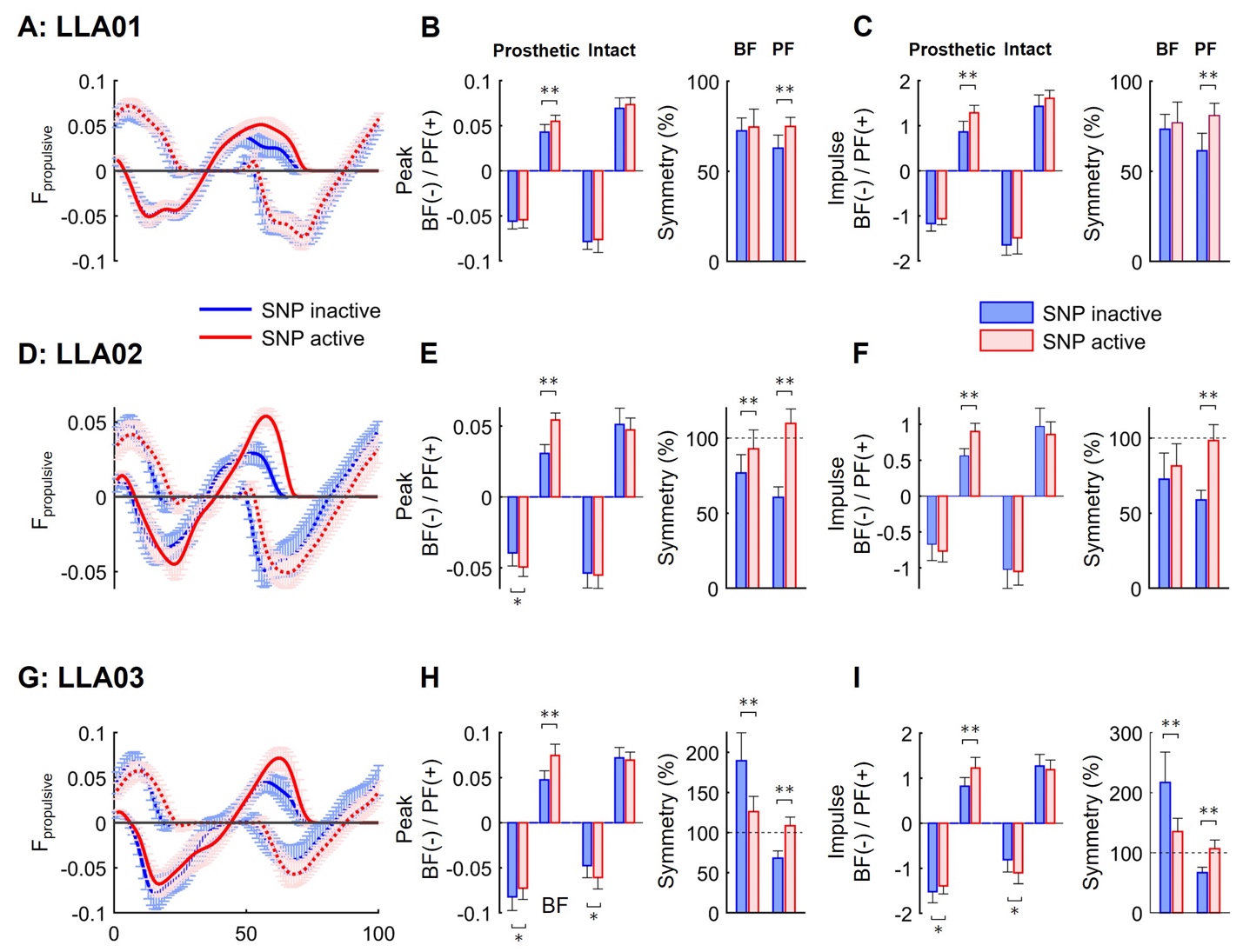

**Fig. S3. Individual SNP users’ anteroposterior ground reaction force (GRF) measures. Related to Fig. 3A.** Mean normalized anterior (propulsive) force (F_propulsive_) and posterior (braking) force (F_braking_) from prosthetic (solid line) and intact (dotted line) limbs are plotted over the prosthetic limb gait cycle for LLA01**(A)**, LLA02 **(D)**, and LLA03 **(G)**. Mean peaks of F_braking_ and F_propulsive_ from each limb and its symmetry between limbs are compared between SNP inactive and SNP active modes for LLA01 **(B)**, LLA02 **(E)**, and LLA03 **(H)**, respectively. In addition, mean impulses of F_braking_ and F_propulsive_ from each limb and symmetry between limbs are compared between SNP conditions for LLA01 **(C)**, LLA02 **(F)**, and LLA03 **(I)**, respectively. Error bars indicate the standard deviation. The asterisk indicates significant differences. ***P*<0.01; **P*<0.05.

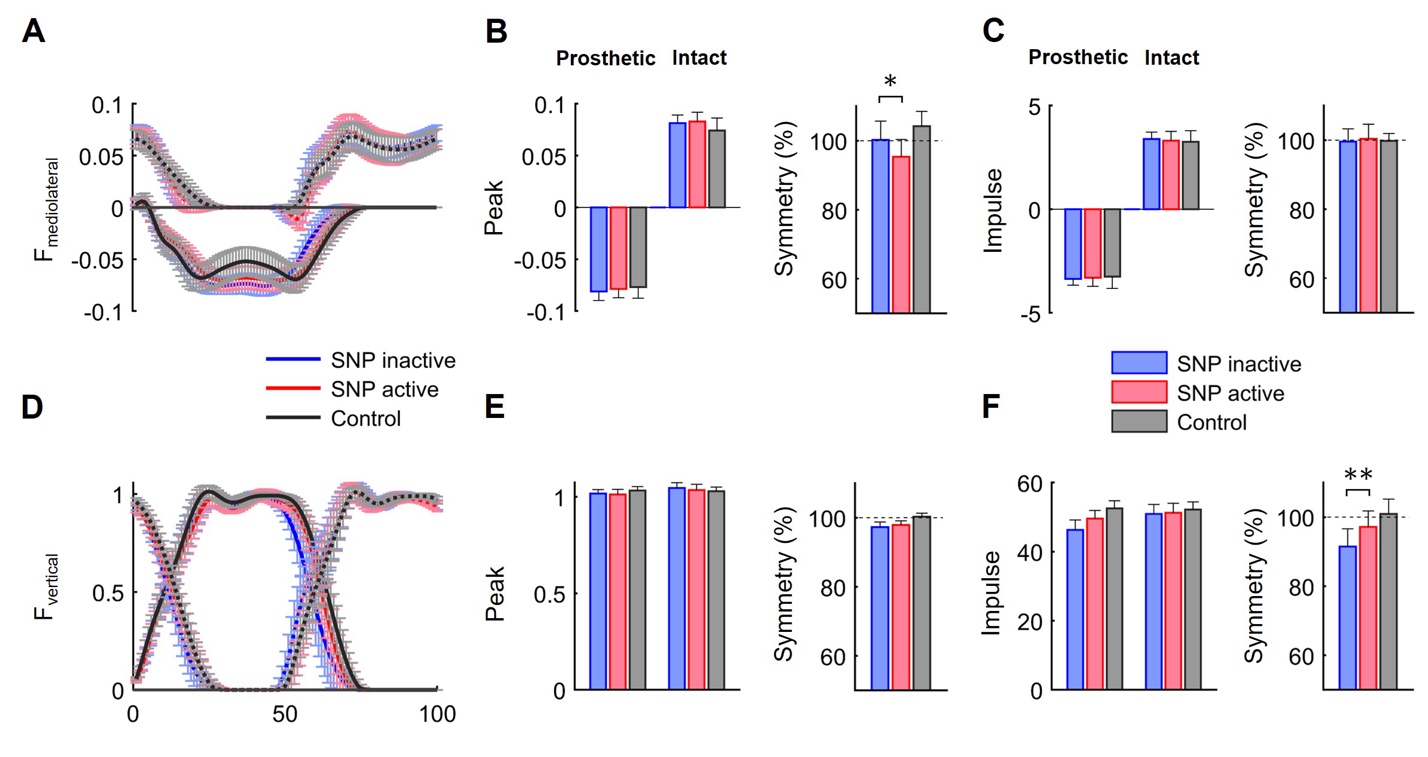

**Fig. S4. GRFs in the mediolateral (F_mediolateral_) and vertical (F_vertical_) directions.** Mean normalized F_mediolateral_ **(A)** and F_vertical_ **(D)** are plotted over the prosthetic limb gait cycle between SNP conditions. The peak values from each limb and symmetry between limbs are shown for F_mediolateral_ **(B)** and F_vertical_ **(E)**. The impulses from each limb and its symmetry between limbs are shown for F_mediolateral_ **(C)** and F_vertical_ **(F)**. Error bars indicate the standard deviation. The asterisk indicates significant differences. ***P*<0.01; **P*<0.05.

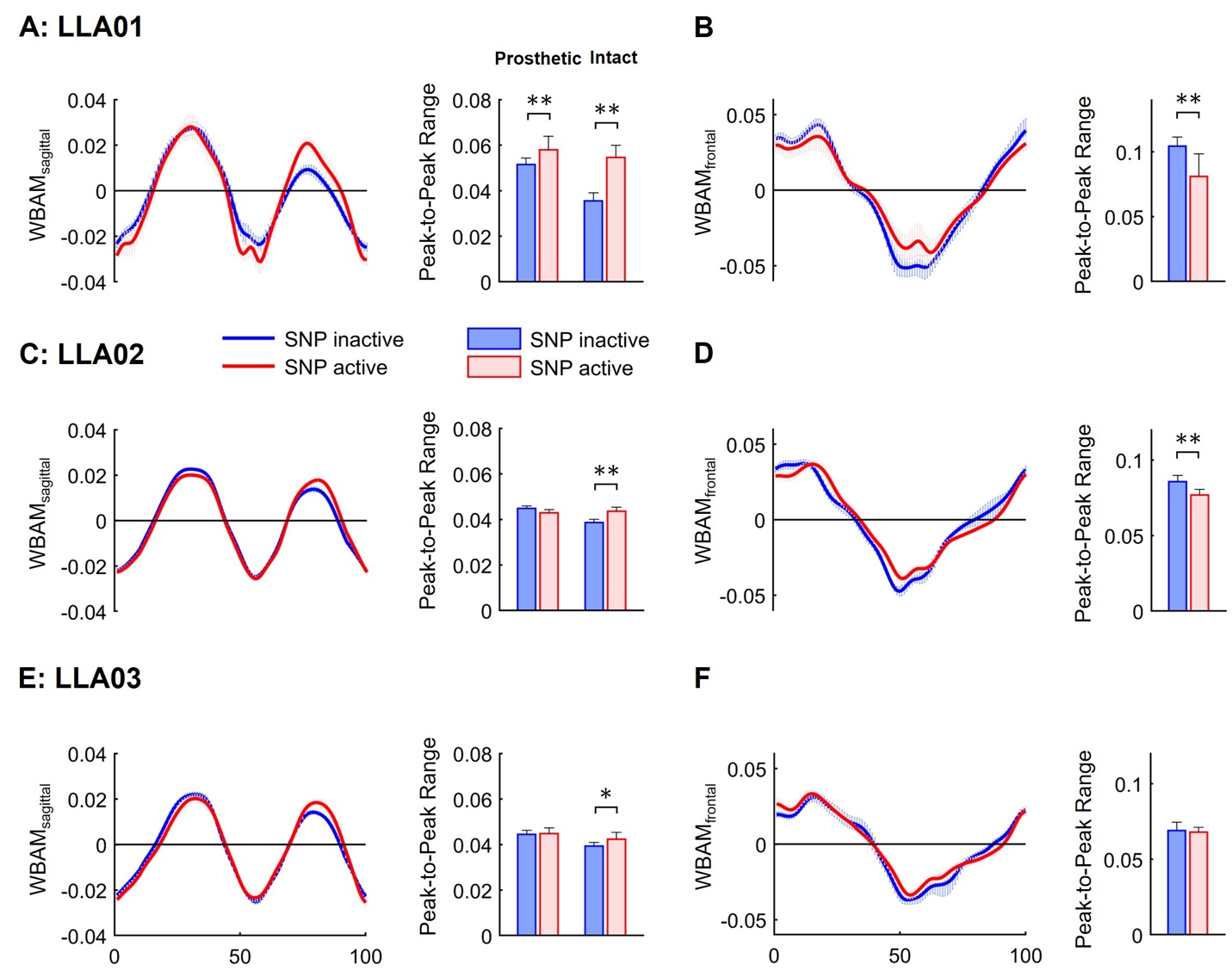

**Fig. S5. Individual SNP users’ WBAM measures. Related to Fig. 3B & C.** Mean normalized WBAM in the sagittal (WBAM_sagittal_ for LLA01 **(A)**, LLA02 **(C)**, and LLA03 **(E)**, respectively) and frontal (WBAM_frontal_ for LLA01 **(B)**, LLA02 **(D)**, and LLA03 **(F)**, respectively) planes are plotted over the prosthetic limb gait cycle between SNP inactive and SNP active modes. The bar graph (next to the line graph) indicates the mean peak-to-peak ranges of WBAM_sagittal_ and WBAM_frontal_ for individual SNP users. Error bars indicate the standard deviation. The asterisks indicate significant differences. ***P*<0.01; **P*<0.05.

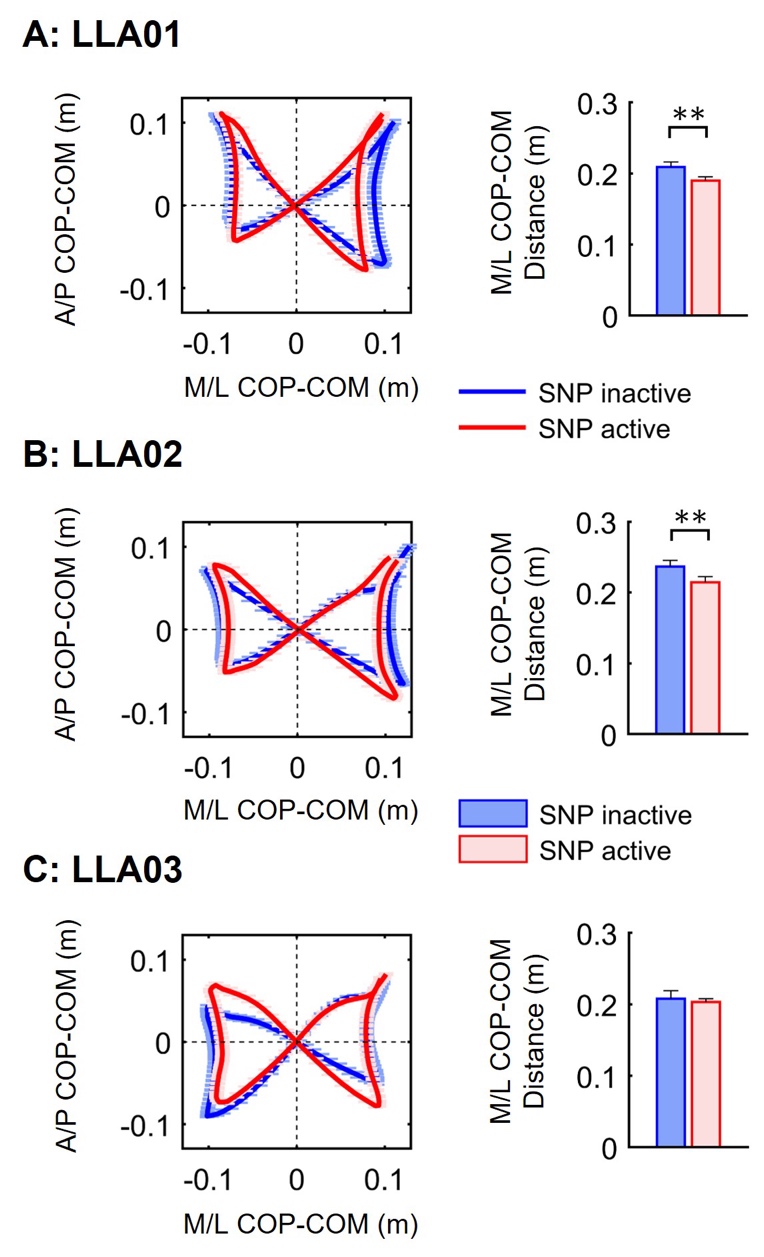

**Fig. S6. Individual COP profile. Related to Fig. 3D.** The line plots show individual SNP user's mean COP trajectories relative to the body’s COM (i.e., COP-COM distance) over the gait cycle for LLA01 **(A)**, LLA02 **(B)**, and LLA03 **(C)**, respectively. The bar graphs indicate the mean COP-COM distance in the mediolateral (M/L) direction between SNP inactive and active conditions. Error bars indicate the standard deviation. The asterisks indicate significant differences. ***P*<0.01.

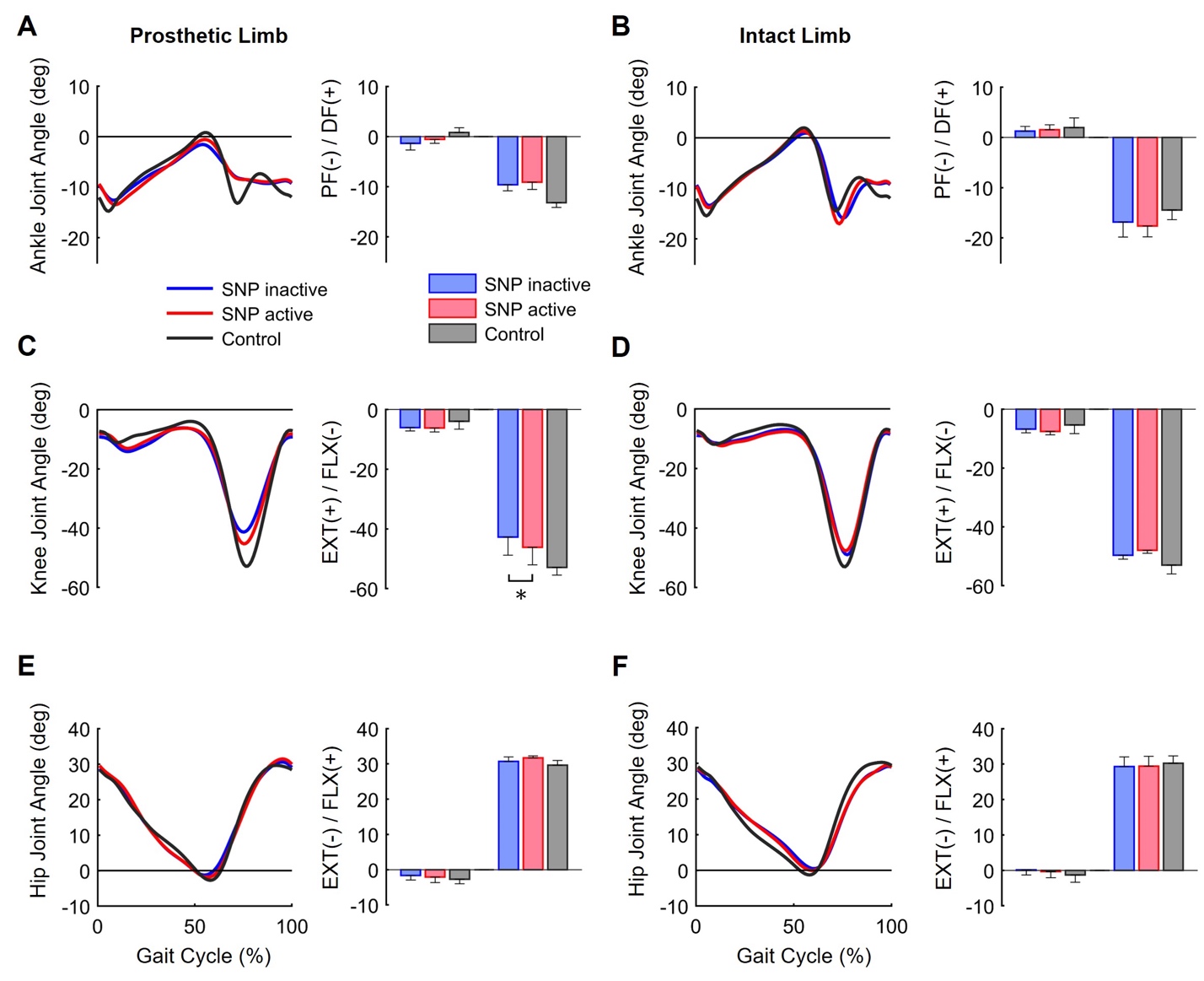

**Fig. S7. Lower-limb joint angles.** Mean ankle plantar-flexion (PF, -) and dorsi-flexion (DF, +), knee flexion (FLX, -) and extension (EXT, +), and hip flexion (FLX, +) and extension (EXT, -) angles are plotted over the prosthetic limb gait cycle for the prosthetic side **(A, C, & E)** and the intact side **(B, D, & F)** between LLAs with SNP inactive (blue), LLAs with SNP active (red), and able-bodied controls (black). Peak values are shown in the bar graphs, respectively. Error bars indicate the standard deviation. The asterisks indicate significant differences. **P*<0.05.

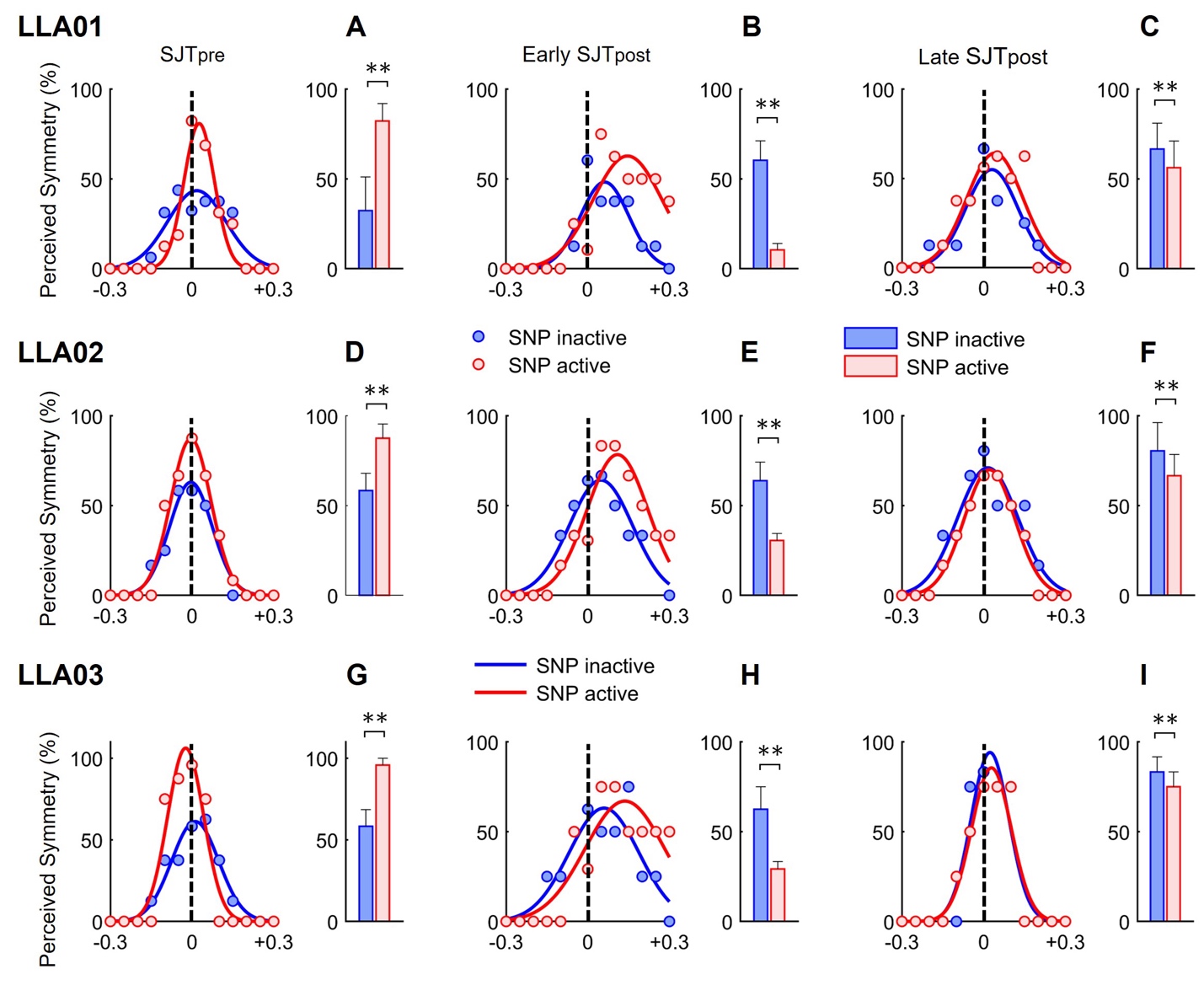

**Fig. S8. Individual SNP users' perceived symmetry during SJT before and after MAT. Related to Fig. 4B, C & Fig. 5E. The dot graphs show the mean perceived symmetry responses across 13 different speed ratios. The bar graphs show the differences in the mean percentages of perceived symmetry responses (when belt speeds were the same) between SNP inactive and active conditions, 1) during the SJT_pre_ (24 trials) for LLA01 (A), LLA02 (D), and LLA03 (G), 2) during the early SJT_post_ (first 12 trials) for LLA01 (B), LLA02 (E), and LLA03 (H), and 3) during the late SJT_post_ (last 12 trials) for LLA01 (C), LLA02 (F), and LLA03 (I), respectively. Error bars indicate the standard deviation. The asterisks indicate significant differences. ***P*<0.01.**

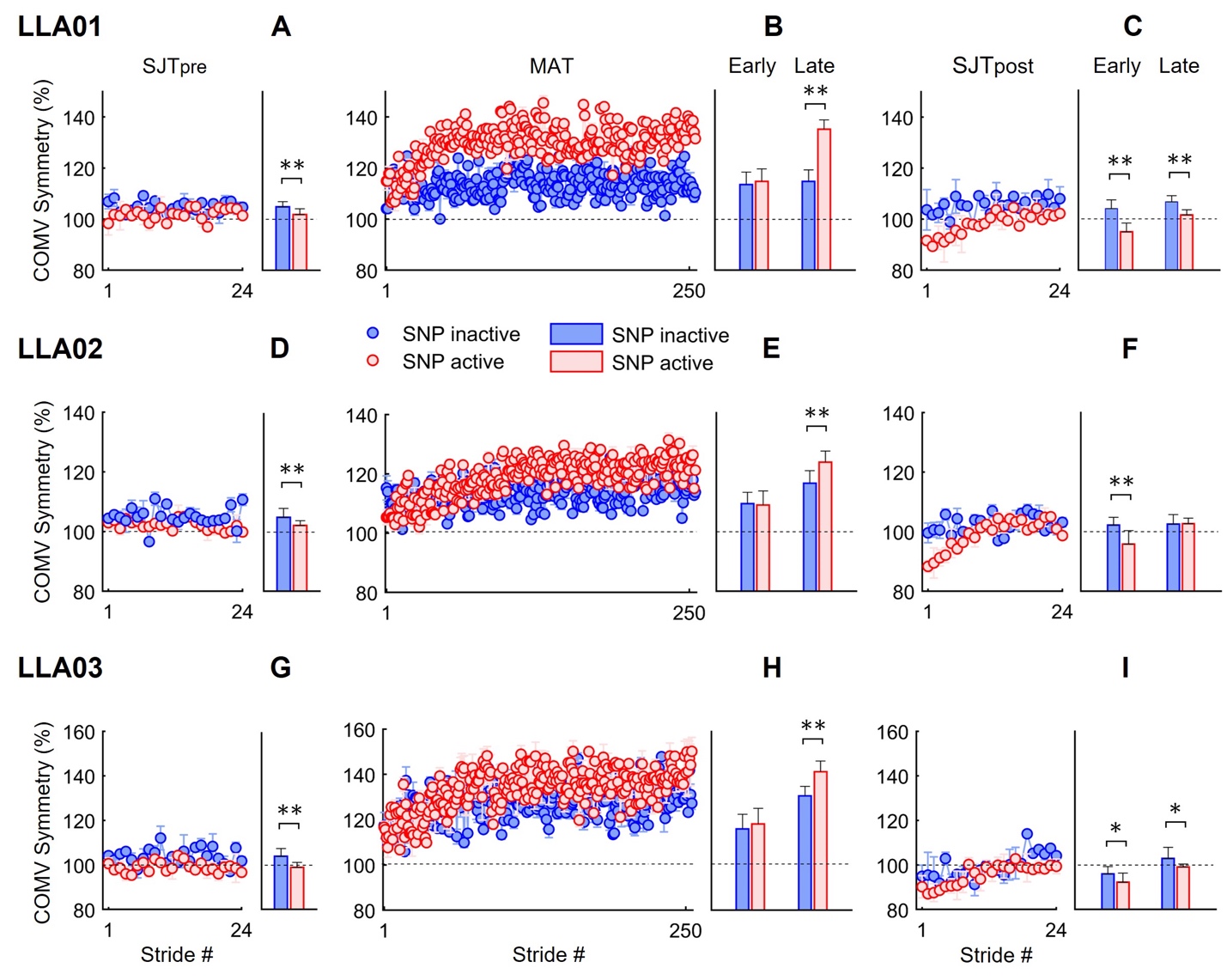

**Fig. S9. Individual SNP users' COM velocity (COMV) symmetry throughout experiments. Related to Fig. 4E, F & Fig. 5B-D. The bar graphs show the differences in the mean COMV symmetry between SNP inactive and active conditions, 1) during the SJT_pre_ (24 trials) for LLA01 (A), LLA02 (D), and LLA03 (G), 2) during the early MAT (first 20 trials) and late MAT (last 20 strides) for LLA01 (B), LLA02 (E), and LLA03 (H), and 3) during the early SJT_post_ (first 12 trials) and late SJT_post_ (last 12 trials) for LLA01 (C), LLA02 (F), and LLA03 (I), respectively. Error bars indicate the standard deviation. The asterisks indicate significant differences. ***P*<0.01; **P*<0.05.**

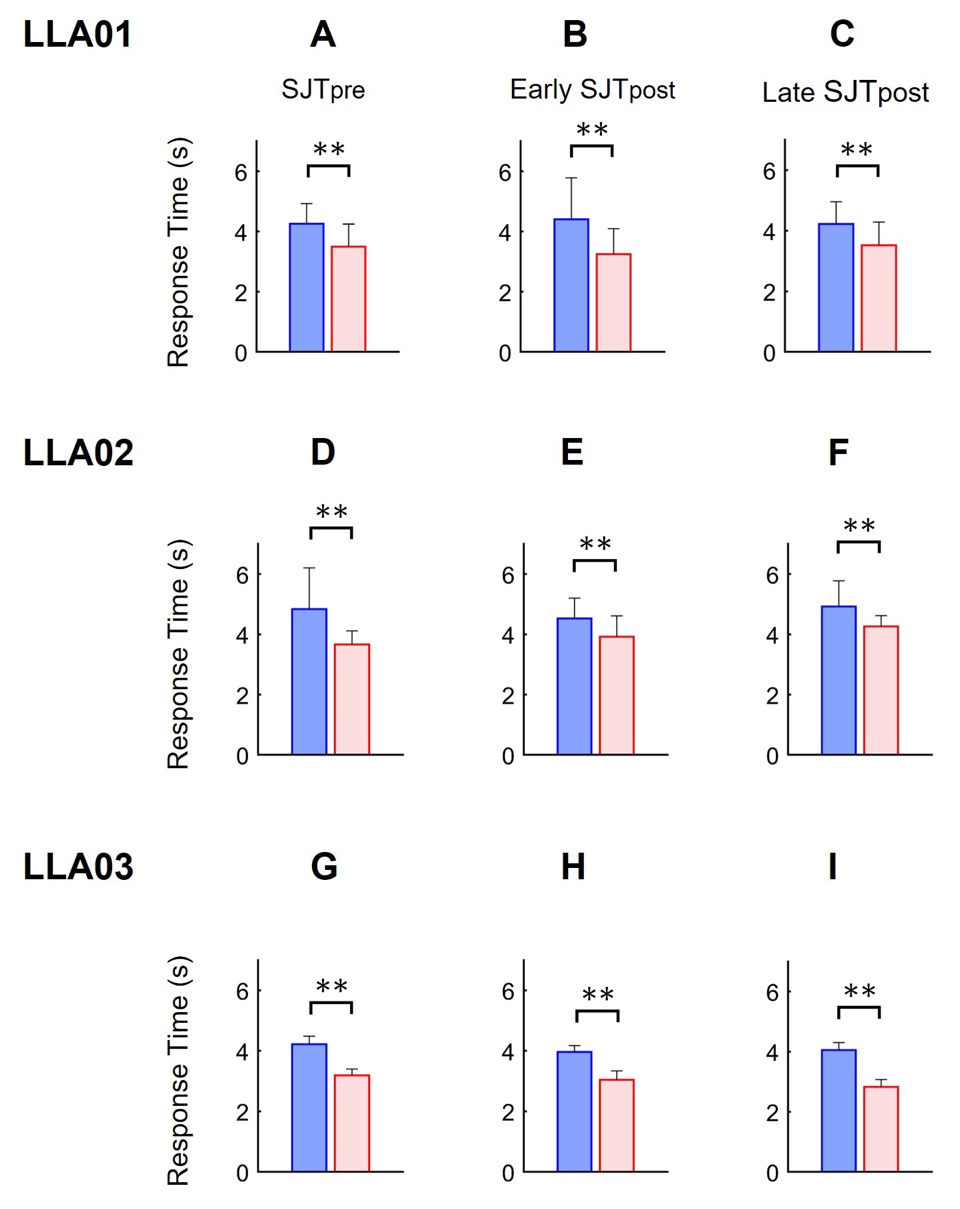

**Fig. S10. Individual SNP users' response time during SJT before and after MAT. Related to Fig. 6A-C. The bar graphs show the differences in the mean response time between SNP inactive and active conditions, 1) during the SJT_pre_ (24 trials) for LLA01 (A), LLA02 (D), and LLA03 (G), 2) during the early MAT (first 20 trials) and late MAT (last 20 strides) for LLA01 (B), LLA02 (E), and LLA03 (H), and 3) during the early SJT_post_ (first 12 trials) and late SJT_post_ (last 12 trials) for LLA01 (C), LLA02 (F), and LLA03 (I), respectively. Error bars indicate the standard deviation. The asterisks indicate significant differences. ***P*<0.01.**
